# Antibody Responses In Non-Severe SARS-CoV-2 Infections Are Driven By CD4+ T cells and Age

**DOI:** 10.1101/2022.04.22.22274032

**Authors:** Amelie E. Murrell, Ewono Eyoh, Jeffrey G. Shaffer, Monika L. Dietrich, Ivy V. Trinh, Thomas J. Yockachonis, Shuangyi Bai, Crystal Y. Zheng, Celia V. Mayne, Sofia E. Cabrera, Anyssa Aviles-Amaro, Addison E. Stone, Saraswatie Rambaran, Sruti Chandra, Debra H. Elliott, Ashley R. Smira, Sara N. Harris, Katharine E. Olson, Samantha J. Bilton, Medea J. Gabriel, Nicole D. Falgout, Emily J. Engel, Alisha D. Prystowsky, Bo Ning, Tony Hu, Jay K. Kolls, Samuel J. Landry, Stacy S. Drury, John S. Schieffelin, Kevin J. Zwezdaryk, James E. Robinson, Bronwyn M. Gunn, Elizabeth B. Norton

## Abstract

SARS-CoV-2 infection causes a spectrum of clinical outcomes and diverse memory responses. Population studies indicate that viral neutralizing antibody responses are protective, but do not always develop post-infection. Other antiviral antibody effector functions, T-cell responses, or immunity to seasonal coronaviruses (OC43, 229E) have been implicated but not defined in all ages. Here, we identify that children and adult subjects generate polyfunctional antibodies to the spike protein after asymptomatic infection or mild disease, with some subjects developing cellular responses without seroconversion. Diversity in immunity was explained by two clusters distinguished by CD4+ T-cell cytokines, age, and antibodies to seasonal coronaviruses. Post-vaccination neutralizing responses were predicted by specific post-infection immune measures, including IL-2, spike-IgA, OC43-IgG1, 229E-IgM. We confirm a key role for CD4+ T cell cytokines in functionality of anti-spike antibodies, and show that antibody diversity is impacted by age, Th/Th2 cytokine biases, and antibody isotypes to SARS-CoV-2 and seasonal coronaviruses.

## INTRODUCTION

Since the start of the coronavirus disease pandemic in 2019 (COVID-19), a central question is what characterizes protective immune responses to the SARS-CoV-2 virus. The risk of SARS-CoV-2 reinfection is estimated as 0–19.5% for at least 10 months following primary infection, indicating that some level of protective immunity develops after infection [1]. Human [2-5] and animal studies [6, 7] have shown that B-cells, antibodies, CD4+ and CD8+ T-cells play essential roles in protection and memory responses. The quality and quantity of these responses are likely key to long-term protective immunity, yet the underlying mechanisms driving these immune responses are still unclear.

Age is a major driver of disease outcomes and significantly impacts the severity and development of SARS-CoV-immunity [8-14]. Children develop functional cellular and humoral immunity to infection but are more likely to be asymptomatic or exhibit mild COVID-19 than adults [8, 10, 15]. Increased age, disease severity, and male sex have all been linked to the development of higher levels of viral-specific IgG and neutralizing antibodies post-infection [16-18]. Population-based studies have shown a central role for these neutralizing antibodies in infection and vaccine-mediated protection to SARS-CoV-2 viral strains [19-21]. Yet, antibody responses exhibit significant variation by subject, and specific protective antibody levels or tests have yet to be identified [22]. In addition, several studies have shown that antibodies with polyfunctional activities that neutralize viral entry and recruit innate immune effector functions, such as complement deposition, neutrophil, and monocyte phagocytosis, and Natural Killer (NK) cell activation, are inversely correlated with disease severity following infection [10, 19, 20, 23]. These reports indicate diversity in SARS-CoV-2 humoral immunity suggesting that the qualitative features of antibodies are likely just as important as titers and neutralizing responses.

T-cell responses also develop early post-infection and are sustained over time. CD4+ T-cell responses are typically characterized by a type 1 phenotype (Th1), with expression of IL-2, IFNγ, and TNFα that are induced early during SARS-CoV-2 infection with a prolonged contraction in patients with severe or mild disease [4, 24-28]. Higher Th1 cell responses have been reported in asymptomatic versus symptomatic patients, but the latter also express more inflammatory cytokines (IL-1β, IL-6, TNFα) [24]. Interestingly, while T-cell responses are linked to the development of antibody responses [3], they are also detected in some people who never developed viral-specific antibodies; a phenomenon termed cellular sensitization without seroconversion [26] that has also been observed with other coronaviruses [29]. CD8+ T-cells have been linked to viral clearance [30, 31], but others have suggested this early control of viremia is from bystander CD8+ T-cells activation and innate immunity [26, 32]. Taken together, these studies indicate SARS-CoV-2-specific CD4+ and CD8+ T-cell responses following infection also vary in magnitude across individuals, but their importance is debated.

Thus, factors contributing to the diversity of SARS-CoV-2 immunity are poorly understood. Notably, most COVID-19 studies have focused on immunity generated following severe infection in hospitalized patients, reflecting less than 10% of infected individuals in the United States. Additional limitations of existing studies include inadequate representation of all age groups for non-severe infections and a focus on antibody effector functions or T-cell immunity, but rarely both. The objective of this study was to address these knowledge gaps by integrating a systems serology approach with T-cell and cytokine measures to generate a comprehensive evaluation of post-infection immunity and longevity. To do so, we compared immune responses as a function of age (1–79 years) in a cohort of infected, non-hospitalized versus non-infected subjects spanning pre- and post-COVID-19 and vaccination periods.

## RESULTS

### Demographics of the Community Seroepidemiology and Immunity (CSI) cohort and symptoms experienced across cohort subjects and households

We recruited a cohort of 91 individuals from 45 households located in Greater New Orleans, Louisiana, community from June 2020 to March 2021 (**Table 1)**. Subjects self-reported with suspected or confirmed SARS-CoV-2 infection or exposure between March 2020 and December 2020 before the circulation of Delta or Omicron variants. Each participant completed a survey that collected parameters of demographics, history of infection or exposure, and clinical signs and symptoms (**Table S1**). Importantly, none of the participants had been hospitalized for infection, although ten subjects reported visiting the emergency room for their illness.

**Table 1.** Composition of study cohort by SARS-CoV-2 infection status.

|  |  | SARS-CoV-2 infected |  | SARS-CoV-2 non-infected |  | Total |  |  |  |
| --- | --- | --- | --- | --- | --- | --- | --- | --- | --- |
| Demographic Characteristics | Categories | n=67 |  | n=24 |  | n=91 |  | P value |  |
| Dates of first visit |  | 6/12/2020 - 2/27/21 |  | 6/12/2020 - 12/15/20 |  | 6/12/2020 - 2/27/21 |  | Pearson chi square | Missing values |
| Households, No. |  | 35 |  | 12 |  | 45 |  | 0.0006 |  |
| Subjects with follow-up visit, No. (%) |  | 21 | 31.3% | 1 | 4.2% | 22 | 24.2% | 0.0269 |  |
| Age, Median (Range) |  | 31 | 4 - 79 | 34 | 1 - 69 | 31 | 1 - 79 | 0.0147 |  |
|  | <18 years, No. (%) | 7 | 10.4% | 7 | 29.2% | 14 | 15.4% | 0.014 |  |
|  | 18-50 years, No. (%) | 37 | 55.2% | 15 | 20.8% | 52 | 57.1% |  |  |
|  | 51+ years, No. (%) | 23 | 34.3% | 2 | 8.3% | 25 | 27.5% |  |  |
| Female No. (%) |  | 47 | 70.1% | 12 | 50.0% | 59 | 64.8% | 0.0761 |  |
| Ethnicity, No. (%) | White | 56 | 83.6% | 21 | 87.5% | 77 | 84.6% | 0.851 |  |
|  | Other | 11 | 16.4% | 3 | 12.5% | 14 | 15.4% |  |  |
| Body Mass Index Category, No. (%) |  |  |  |  |  |  |  |  |  |
|  | Underweight | 3 | 4.5% | 2 | 8.7% | 5 | 5.6% | 0.2506 | 2 |
|  | Healthy Weight | 39 | 59.1% | 8 | 34.8% | 47 | 52.8% |  |  |
|  | Overweight | 15 | 22.7% | 8 | 34.8% | 23 | 25.8% |  |  |
|  | Obese | 9 | 13.6% | 5 | 21.7% | 14 | 15.7% |  |  |
| Smoking, No. (%) |  | 9 | 13.6% | 0 | 0.0% | 9 | 10.0% | 0.0565 | 1 |
| Has a chronic conditions, No. (%) |  | 19 | 30.6% | 7 | 31.8% | 26 | 31.0% | 0.9186 | 7 |
|  | Hypertension, No (%) | 6 | 9.7% | 2 | 9.1% | 8 | 9.5% | 0.9358 | 7 |
|  | Asthma, No. (%) | 3 | 4.8% | 2 | 9.1% | 5 | 6.0% | 0.4689 | 7 |
|  | High cholesterol, No. (%) | 9 | 14.5% | 1 | 4.5% | 10 | 11.9% | 0.2147 | 7 |
| **COVID specific details** |  |  |  |  |  |  |  |  |  |
| Reported symptoms with illness, No. (%) |  | 58 | 86.6% | 14 | 58.3% | 72 | 79.1% | 0.0035 |  |
| visited the ER for suspected illness, No. (%) |  | 9 | 13.4% | 1 | 4.2% | 10 | 11.0% | 0.213 |  |
| tested SARS-CoV-2+ (PCR self), No. (%) |  | 42 | 62.7% | 0 | 0.0% | 42 | 46.2% | <0.0001 |  |
| tested SARS-CoV-2+ (PCR household), No. (%) |  | 53 | 79.1% | 0 | 0.0% | 53 | 58.2% | <0.0001 |  |
| Plasma viral RNA detected, No. (%) |  | 11 | 16% | 0 | 0% | 0 | 12% | 0.0343 |  |
| Serostatus, No. (%) | anti-S positive | 54 | 80.6% | 0 | 0.0% | 54 | 59.3% | <0.0001 |  |
|  | anti-S indeterminant | 6 | 9.0% | 3 | 12.5% | 9 | 9.9% |  |  |
|  | anti-S negative | 7 | 10.4% | 21 | 87.5% | 28 | 30.8% |  |  |
|  | anti-N positive | 32 | 47.8% | 0 | 0.0% | 32 | 35.2% | <0.0001 |  |
|  | anti-N indeterminant | 28 | 41.8% | 3 | 12.5% | 31 | 34.1% |  |  |
|  | anti-N negative | 7 | 10.4% | 21 | 87.5% | 28 | 30.8% |  |  |
| met COVID-19 case definition |  | 51 | 76% | 0 | 0% | 51 | 56% | <0.0001 |  |
| mean days from possible exposure (range) |  | 93 | (23-252) | 133 | (10-289) | 104 | (10-289) | 0.1119 |  |

SARS-CoV-2 infection in 67 subjects (74%) was confirmed either by a PCR test (n=42), SARS-CoV-2 spike or nucleoprotein specific IgG by ELISA and Luminex testing (54 and 32 subjects, respectively) or viral RNA detection in plasma (n=11) by a sensitive CRISPR assay [33, 34]. The remaining 24 subjects exhibited no evidence of SARS-CoV-2 infection. Blood samples were collected at a range of times post-symptom onset or days post-exposure (**Figure 1A**) (range of 10–289 days; median of 99 days from subjects), and subjects exhibited a range of disease severity irrespective of household (**Figure 1B**).

**Figure 1.**
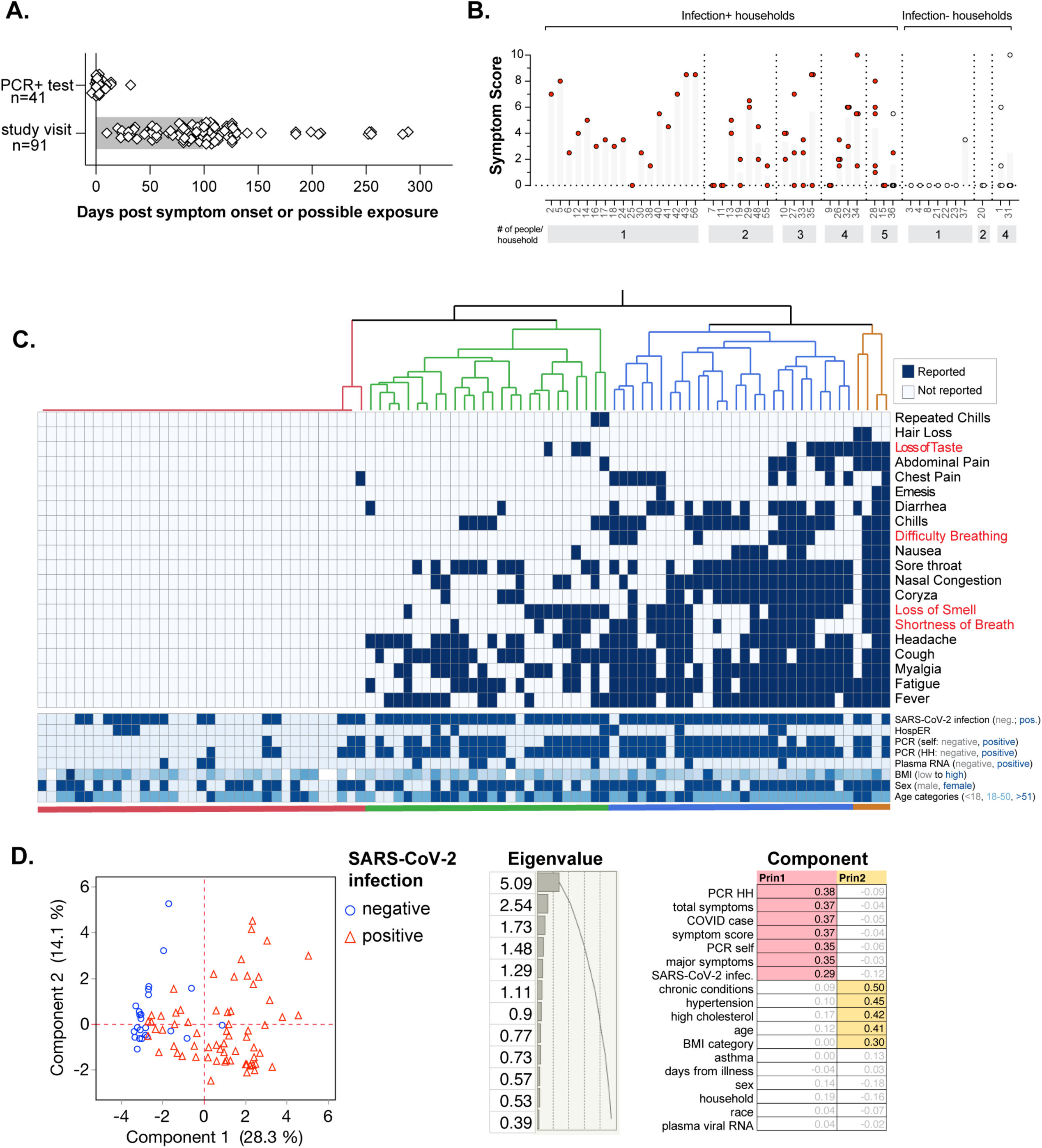
Cohort composition includes a range of asymptomatic or mild to moderate disease subjects. (A) Days post-symptom onset or possible viral exposure for last SARS-CoV-2 PCR positive test for each subject and indicated day of first study visit. The bars represent the median number of days. (B) Symptom score as a composite of major and minor COVID-19 symptoms, range 0–21. Red circles indicate infected subjects and white circles indicate uninfected subjects. (C) Hierarchical clustering of individual subjects by symptoms with associated heatmap for reported symptom or infection and demographic information listed underneath (blue colors or color ranges indicate positive, high, female, or older age categories). Major symptoms are shown colored red. (D) PCA analysis of subject’s infection and demographic data colored by SARS-CoV-2 infection status with eigenvalues and eigenvector variables shown for components 1 and 2.

Hierarchical clustering analysis on reported symptoms demonstrated that five major symptom clusters were observed across the cohort (**Figure 1C**), including no symptoms, and symptoms known to track with severe disease such as difficulty breathing, shortness of breath, and loss of taste/smell. To identify those demographic parameters associated with infection status, principal component analyses (PCA) were performed, where components were ordered according to their proportion of explained variability in the data. PCA identified two components that explained at least 10% of the variability in the data (**Figure 1D**). Infected and uninfected individuals were distinguished in Component 1 by positive PCR tests and variables related to reporting symptoms for SARS-CoV-2 infection. Select demographic variables further characterized individuals across Component 2, including subject age, chronic conditions (e.g., hypertension, high cholesterol), and BMI, but not sex, race, or household. These analyses demonstrate that medical co-morbidities observed in hospitalized COVID-19 patients were also relevant in this non-hospitalized cohort.

### Polyfunctional humoral immunity is induced following SARS-CoV-2 infection

Given the central role of antibodies in SARS-CoV-2 immunity, we quantitatively and qualitatively profiled SARS-CoV-2 specific humoral immunity in the cohort. Using multiplexed Luminex analyses, we measured plasma levels of different IgG subclasses (IgG1, IgG2, IgG3, IgG4), IgA (IgA1, IgA2), and IgM-specific for SARS-CoV-2 antigens (full-length spike protein, the receptor-binding domain (RBD), and the nucleoprotein [N]) and seasonal coronaviruses (OC43, HKU1, 229E, and NL63) spike proteins.

As expected, SARS-CoV-2 specific antibodies were significantly elevated in subjects with SARS-CoV-2 infection compared with uninfected individuals (**Figure 2A, Figure S1**). No significant differences were observed between infection groups with respect to the levels of antibodies against the seasonal coronaviruses. Sampling times post-infection differed across the cohort. While the time between sampling and infection did not correlate with increasing/decreasing IgG1 across individuals, elevated levels of IgG4 were observed in samples collected further from infection, suggesting temporal development of virus-specific IgG4 (**Figure 2B**). A subset of subjects (n=18) returned for follow-up visits six months after visit 1 (and before any vaccination). In these subjects, spike- and N-specific IgG responses were relatively stable over time (Figure S2). However, a small but significant increase in spike-specific IgA1 was observed in the follow-up visit (**Figure S2**).

**Figure 2.**
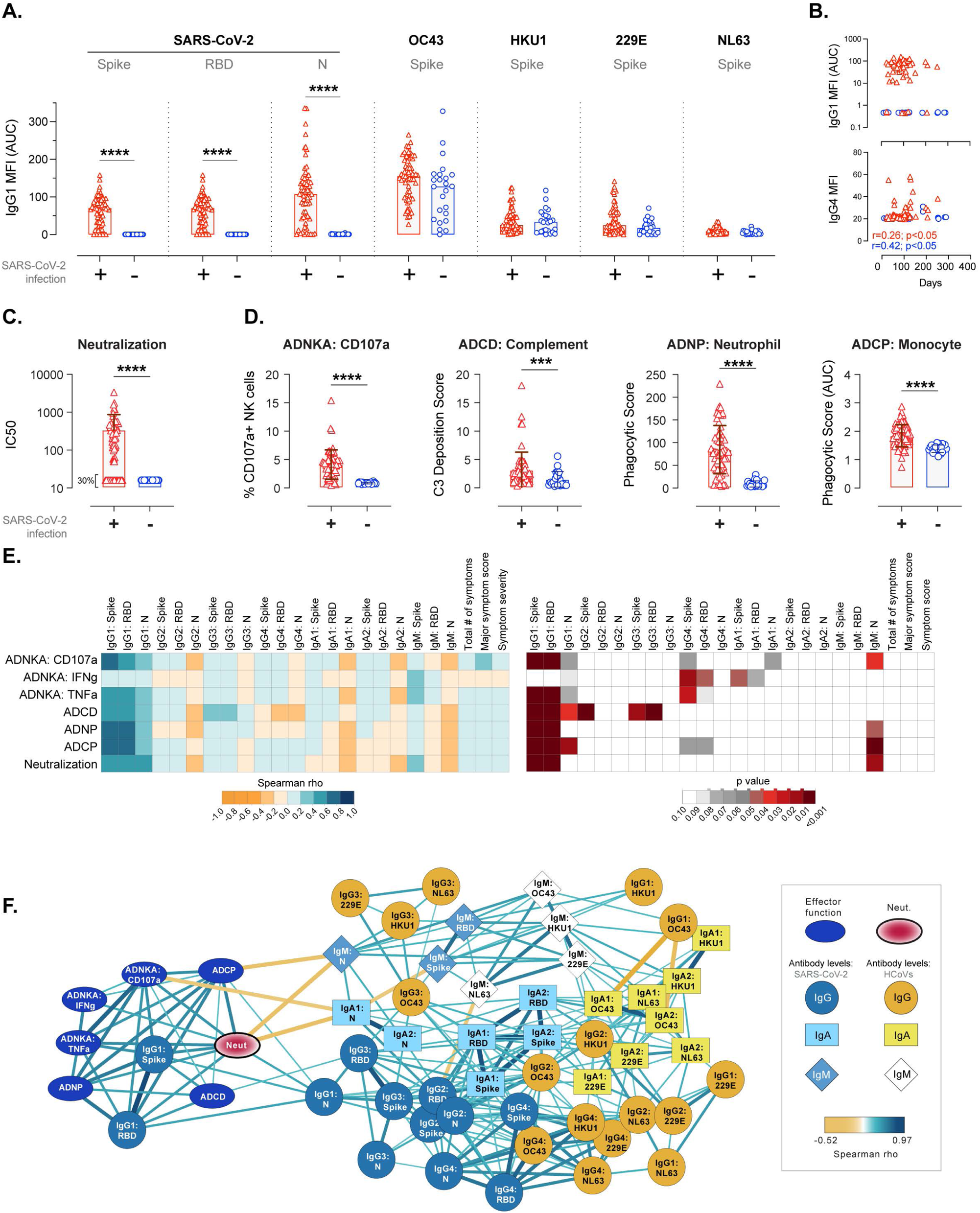
Antibody isotypes dictate effector functions post-infection. (A) IgG1 responses to spike, RBD or N SARS-CoV-2 specific antigens and spike-specific responses to seasonal coronaviruses OC43, HKU1, 229E, NL63 for each subject by SARS-CoV-2 infection status. (B) IgG1 or IgG4 spike-specific expression by days post-infection or possible exposure with significant Spearman’s correlations indicated. (C) Neutralization responses by SARS-CoV-2 infection status. (D) ADNKA, ADCD, ADNP, or ADCP responses by SARS-CoV-2 infection status. (E) Correlation matrix for indicated measures with heatmaps of Spearman’s rho values (left) and p values (right). (F) Network of indicated antibody responses with the color of connecting lines representing Spearman’s rho values. Bars represent mean +/- SD. Significance by Mann Whitney indicated as p<0.05 (*), <0.01 (**), or <0.001 (***). Area under the curve (AUC).

As both neutralizing antibodies and induction of antibody-mediated innate immune effector functions have been associated with antibody-mediated protection against SARS-CoV-2 infection [10, 20, 21], the ability of each subject’s plasma to neutralize SARS-CoV-2 D614G pseudoviruses and induce innate immune effector functions against spike-coated targets (phagocytosis by monocytes (ADCP) or neutrophils (ADNP), complement deposition (ADCD), and NK cell activation (ADNKA) was determined. Plasma from most infected individuals was found to possess neutralizing activity and not be related to time from infection, yet interestingly a sizable subset of people (n=20; 29.8% of infected individuals) did not make neutralizing antibodies (**Figure 2C**). Similar results were observed for induction of innate immune effector functions (**Figure 2D)**, although the profiles among individuals varied (**Figure S3A**). To determine if specific antibody subclasses or isotypes were associated with antiviral effector functions, we performed a correlation analysis between SARS-CoV-2 specific antibody levels and effector functions. Among infected individuals, spike and RBD-specific IgG1 were highly correlated with antiviral functional activity and symptom score (**Figure 2E, Figure S1B**), suggesting that IgG1 predominantly drives these antiviral functions, although it should be noted that IgG3 strongly correlated to ADCD. N-specific IgM was negatively associated with many of these effector functions including ADNKA, ADNP, ADCP, and neutralization. While IgM is often observed early in infections (prior to IgG response), IgM levels were detected that were not significantly associated with days post-infection or inversely related to virus-specific IgG. Thus, the IgM response we detected does not seem to have a temporal relationship with antibody maturation and class-switching. Network analyses of all antibody responses against SARS-CoV-2 and the seasonal coronaviruses further confirmed the association of SARS-CoV-2 spike and RBD-specific IgG1 with qualitative antibody functions but with limited associations to other isotypes and specificities (**Figure 2F**). Together, these data highlight the diversity of qualitative humoral immune responses observed among infected individuals.

### CD4+ and CD8+ T-cell immunity are induced following infection, and CD4+ Th1 responses strongly correlated with SARS-CoV-2 serum IgG and neutralization

To determine cellular immunity post-infection, we evaluated stored PBMCs from all subjects for CD4+ and CD8+ T-cell reactivity and cytokine secretion in response to SARS-CoV-2 spike protein, nucleoprotein, or matrix peptide pools after *in vitro* stimulation for 24h. Activation was assessed by flow cytometry of stimulated versus non-stimulated PBMC cultures by gating for CD134-high expression on CD4+ T cells or CD69 expression on CD8+ T-cells as a modified activation-induced marker expression (AIM) (**Figure S4A,B**). As previously reported in SARS-CoV-2 infected subjects [4, 24, 26], significant CD4+ and CD8+ T cell AIM to spike protein or peptide were observed compared to non-infected subjects (**Figure 3A**). CD4+ T cell AIM responses were stable over 6 months in the individuals who returned for a follow-up visit (**Figure S2E-F)** and were not correlated with the day of sample collection (**Figure S4F**). Interestingly, a subset of infected subjects had detectable T-cell responses with no observable antibody responses (**Figure S3A,B**), indicating that these subjects exhibit cellular sensitization without seroconversion [26].

**Figure 3.**
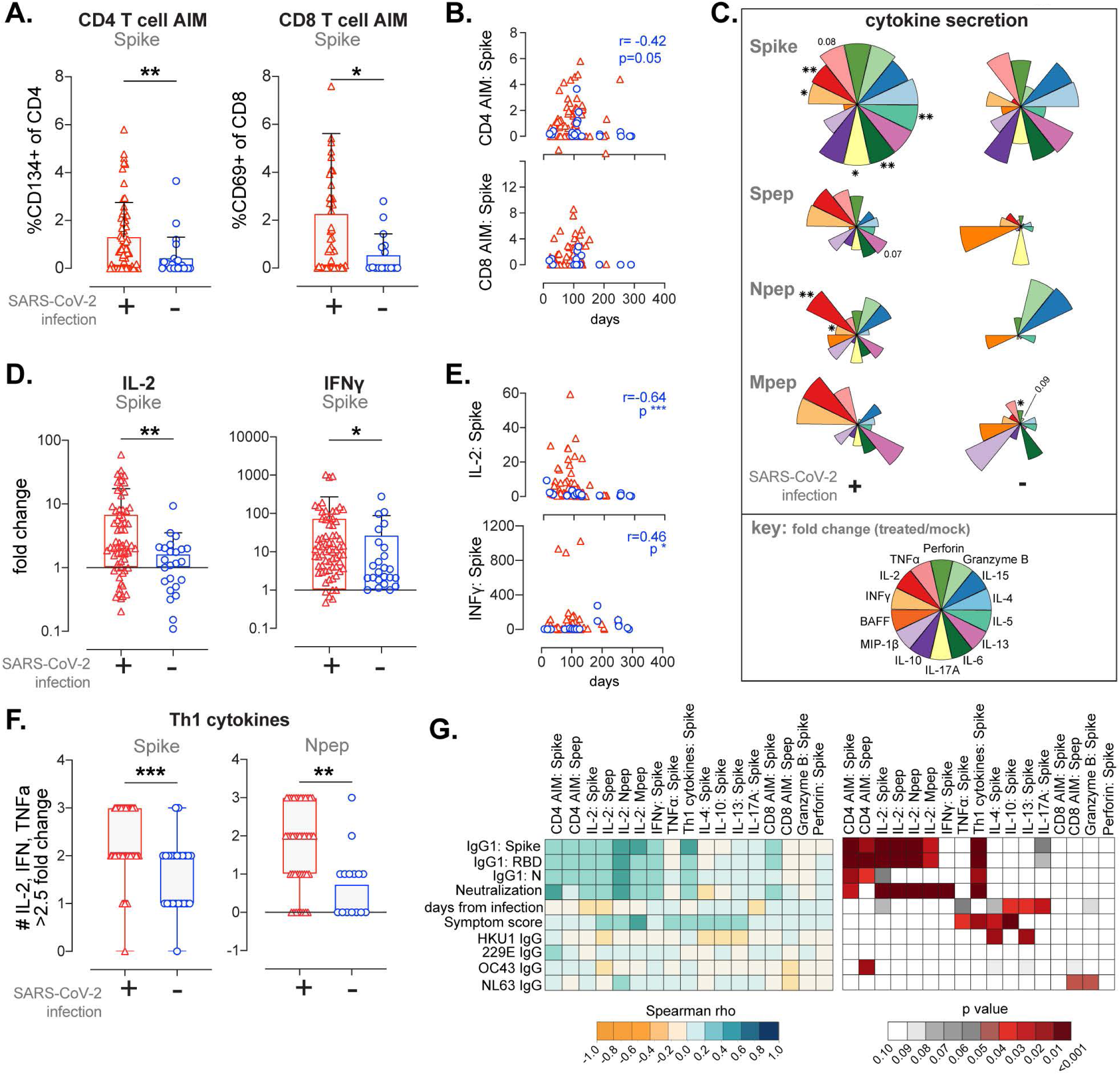
CD4 Th1 responses to spike, nucleoprotein, and matrix antigens strongly correlate with SARS-CoV-2 serum IgG and neutralization responses. (A) Spike-specific AIM changes in CD4 or CD8 T cells by SARS-CoV-2 infection status (% of CD4 or CD8). CD8 T cell AIM excludes three high responders in the SARS-CoV-2 infection group (values of 8.6, 14.7, and 15.6). (B) T cell AIM results by day post-infection or possible exposure with significant Pearson’s correlations indicated. (C) Pie charts of log-transformed mean cytokine levels to spike protein or peptide pools (spike, nucleoprotein or matrix) expressed as fold change from unstimulated with color key indicated. (D) Spike-specific IL-2 and IFNγ cytokines secretion expressed as fold change from unstimulated by infection status. (E) Cytokine changes by day post-infection or possible exposure with significant Pearson’s correlations indicated. (F) Number of IL-2, IFNγ, or TNFα cytokines expressed at fold change >2.5 from unstimulated cells by SARS-CoV-2 infection status. (G) Correlation matrix for indicated measures with heatmaps of indicated Spearmans’ rho (left) and p values (right). Bars shown at mean +/- SD. Significance by Mann Whitney indicated as p<0.05 (*), <0.01 (**), or <0.001 (***).

To further characterize the types of T cell responses that are induced in infected individuals, we measured cytokines indicative of specific T-helper (Th)-responses, cytotoxic T-cell responses, or chemokines in stimulated PBMC culture supernatants compared with unstimulated PBMC. Induced cytokine secretion was highly correlated among almost all restimulation antigens including spike protein, spike peptide, N peptide, or matrix peptide (**Figure S5**). As anticipated [4, 24-28], Th1-biasing IL-2 and IFNγ secretion was observed in response to spike and nucleoprotein, but not matrix viral antigens, in infected subjects. Th1 cytokine secretion did not correlate with the day of sample collection (**Figure 3C-E, Figure S4F**) but declined in follow-up visits of infected subjects (**Figure S2G-J**). Although TNFα secretion alone was not associated with infection, a 2.5- or 4-fold change in the three Th1-associated cytokines (IL-2, IFNγ, and TNFα) induced by spike protein and nucleoprotein peptides was observed in infected subjects (**Figure 3F, Figure S4E, S4G)**. Restimulation with spike protein also induced Th2 and Th17 cytokines, including IL-5 and IL-17, the latter of which was highly correlated to IL-2 responses (**Figure S5**). Significant changes in parameters such as IL-4, IL-10, Granzyme B, and perforin were not observed for any stimulation condition related to infection status (**Figure S4E)**.

As T-cell and cytokine secretion can impact the generation of antibodies and have been implicated in pre-existing immunity to seasonal coronaviruses, we correlated cellular immunity measures with SARS-CoV-2 and seasonal coronavirus IgG antibodies (**Figure 3G**). CD4+ T-cell AIM, IL-2 secretion, and Th1-associated cytokines were significantly correlated to plasma levels of SARS-CoV-2 spike, RBD, and N IgG. Spike peptide pools but not protein for AIM significantly correlated to OC43 and NL63 IgG. In contrast, symptom score was highly associated with TNFα, IL-4, and IL-10, even though these cytokines individually were not different between infected and non-infected subjects, suggesting that these cytokines track with disease severity.

### Age drives immunity to SARS-CoV-2 infection

The diversity of the study cohort (**Table 1**) allowed us to evaluate the relationship between age, sex, and SARS-CoV-2 immunity. In infected subjects, higher levels of chronic conditions and SARS-CoV-2 symptom scores were associated with increasing age (**Figure 4A**) and correlated with increased levels of spike-specific IgG1, neutralizing antibodies, IL-2, CD8+ T-cell activation, and the related antiviral cytokines perforin and Granzyme B (**Figure 4B-D**). Minimal changes in seasonal coronavirus IgG were observed with age (**Figure 4B**). Sex had no significant impact on these measures (**Figure 4B**); however, both sex and age were significantly related to IgM antibody levels to Spike and RBD (**Figure 4B**).

**Figure 4.**
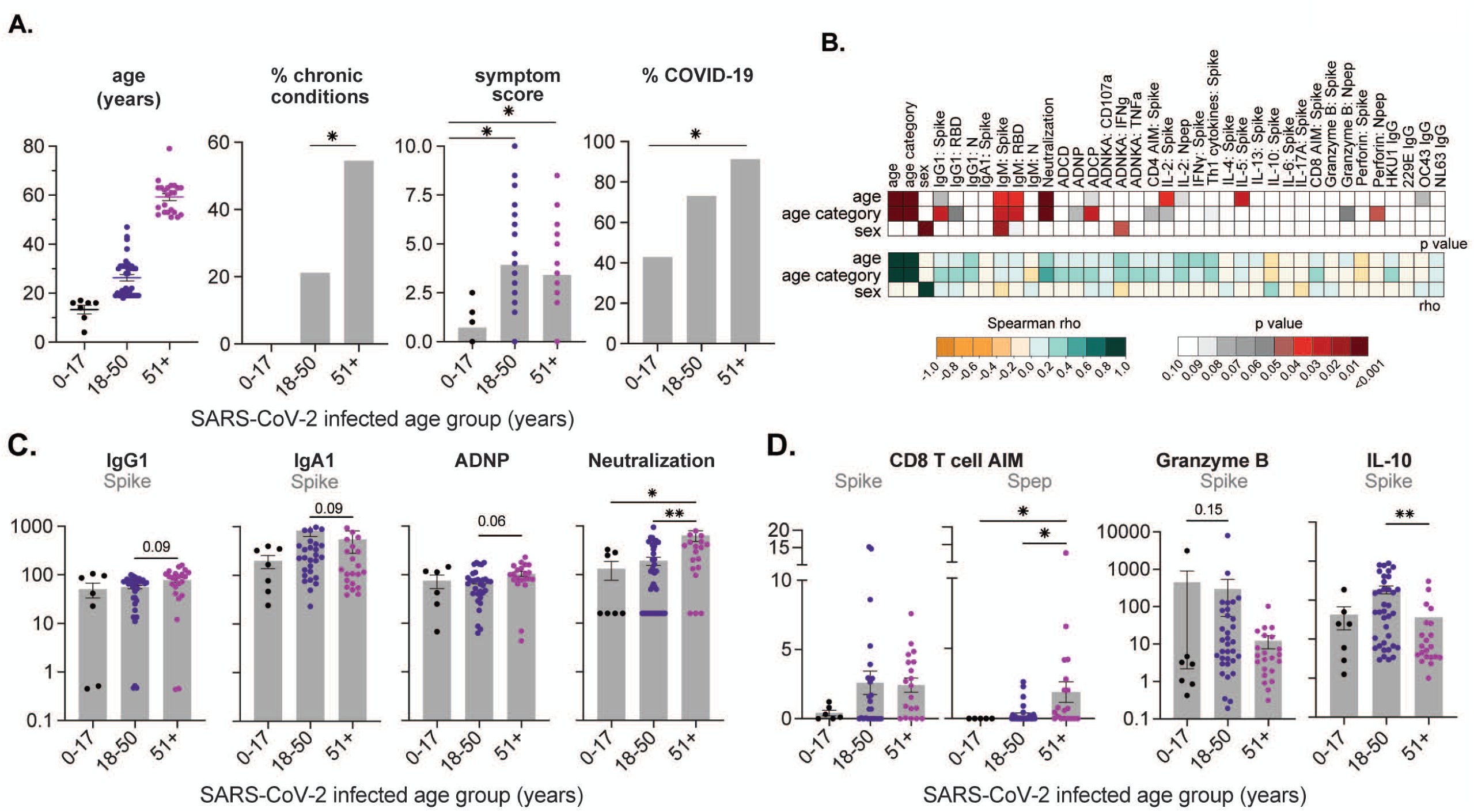
Age is a significant driver of disease severity and development of SARS-CoV-2 immunity. (A) Age of each subject, percent of individuals with chronic conditions, symptom score of each subject and percent of individuals that meet COVID-19 case definition by age category. (B) Correlation matrix for age, age category and sex (male = 1, female = 2) with heat maps of indicated Spearmans’ P or rho values. (C) Spike-specific responses for IgG1 and IgGA1, ADNP, and neutralization responses of each subject by age category. (D) CD8 AIM changes for spike protein or peptide pool and Granzyme B and IL-10 fold change for spike protein by age category. Significance by ANOVA with Dunn’s post-test or logistic regression model with pairwise contrast indicated as P<0.05 (*), <0.01 (**), or <0.001 (***).

### Immunity in infected subjects can be separated into two groups based on Th1 or Th2 biased immune responses and antibody isotypes (IgM/IgG1 or IgG2/IgG4)

To dissect the relationship between cellular and serologic immunity with all disease parameters and demographic information, we used t-SNE and UMAP data visualizations for immune response to SARS-CoV-2 proteins or peptides. These analyses resulted in two distinct populations (e.g., t-SNE 1 and t-SNE 2) verified by k-means clustering (**Figure 5A, Figure S6A**). These populations were compared for all measures of SARS-CoV-2 immunity, demographic, infection, or other seasonal coronavirus variables (**Table S2)**. Across the demographic and disease variables, older age was associated with population 1, and minor symptoms or specific symptoms (nausea, loss of taste, runny nose, nasal congestion) were associated with population 2 (**Figure 5B, Figure S6B,D**). Sex, co-morbidities, infection status, or time from infection was not associated with either population. With regard to immune measures, population 1 was defined by typical SARS-CoV-2 immunity, including IL-2 secretion to spike, CD4+ AIM, antiviral antibody functions (ADCP, ADNP), and elevated levels of IgM to SARS-CoV-2 spike or seasonal coronaviruses (e.g., HKU1, **Figure 5C**). Neutralization was not directly associated with either cluster but correlated strongly with Th1-responses and IL-2 in population 1 (**Figure 5D, Figure S6E**). Alternatively, population 2 was characterized by non-classical immune measures indicative of a Th2-biased response, including IL-4, IL-13 and IL-10 secretion to spike and peptide pools and development of higher levels of IgG2 and IgG4 to RBD (**Figure 5B, Table S2**). Neutralization in this population did not correlate to Th1 cytokines but was negatively correlated with IL-4 (**Figure 5D, Figure S6E**). In addition, perforin and ADNK activities were higher, suggesting population 2 may have higher NK or NKT cell activity. These data indicate that within this non-hospitalized cohort, SARS-CoV-2 immunity can diverge into Th1-biased immunity (observed in population 1) or Th2-biased immunity (observed in population 2), likely related to patient age. It is possible that lingering viral infection or antigen also contribute to population differences, as this showed up in both UMAP and correlation analyses. (**Figure 5E, Figure S6B**).

**Figure 5.**
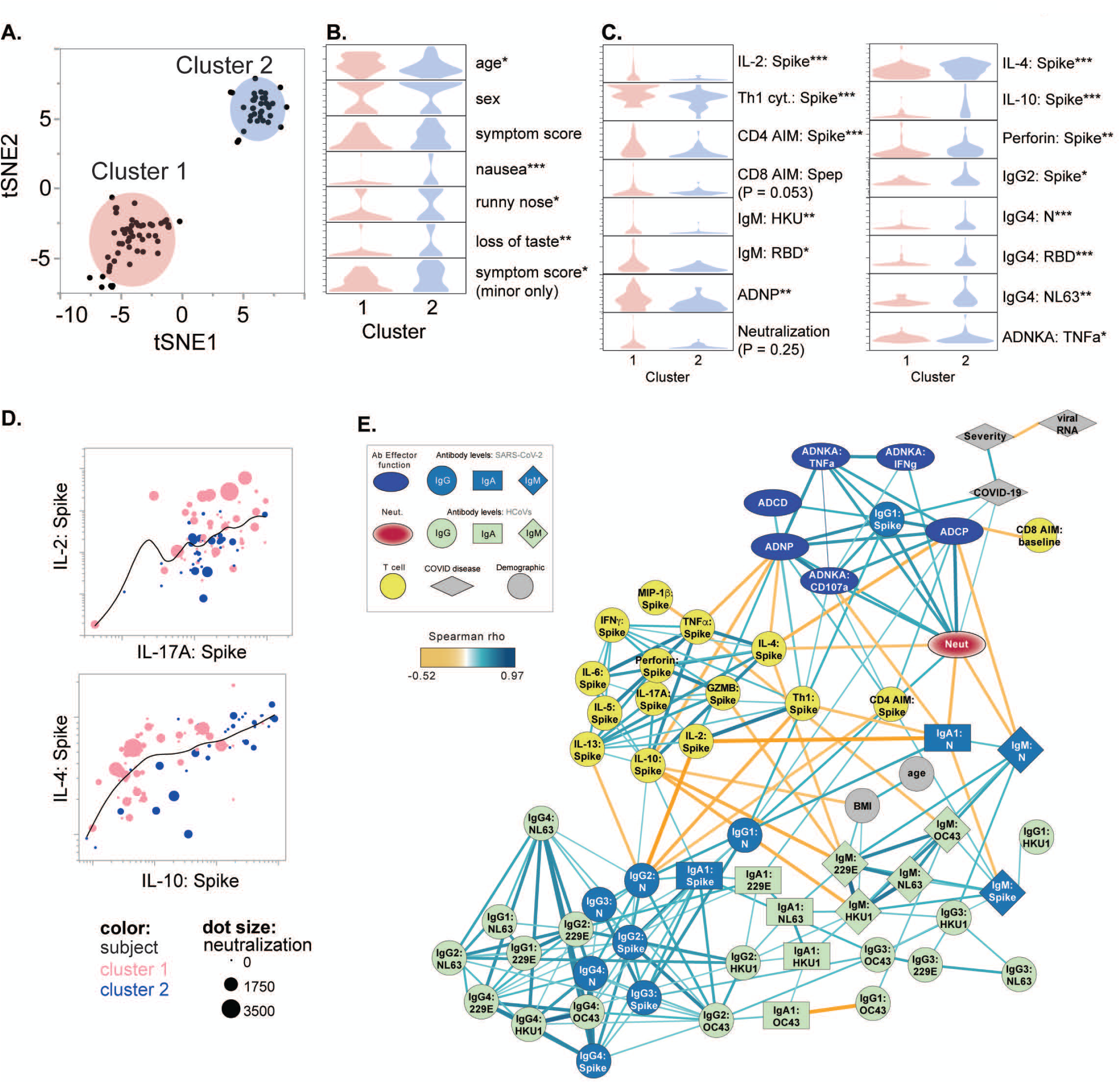
SARS-CoV-2 immunity can be clustered into two populations based on age, Th1/Th2 responses, antibody isotypes, and effector functions. (A) An imputed dataset for all SARS-Co-V immunologic variables was used for t-SNE analysis and then K-means clustering for comparison of variables. No seasonal coronavirus data, infection history or demographic data was used. All imputed data were required to have at least 60% of non-missing cell values. (B) Graphs of demographic and SARS-CoV-2 infection variables defined by cluster. (C) Graphs of SARS-CoV-2 or seasonal coronavirus immunity variables defined by cluster. Significant Kruskal-Wallis test between tSNE clusters indicated by P<0.05 (*), <0.01 (**), or <0.001 (***). (D) Dot plots by indicated measures for subjects included in cluster 1 (pink) or 2 (blue) with spline trend line (black) shown in log scale. (E) Correlation network across immune and disease features that were significantly associated across SARS-CoV-2 infected individuals.

These observations were complemented with a network analysis of immune parameters in SARS-CoV-2 infected subjects, which demonstrated that neutralizing and functional antibody levels were associated with CD4+ T cell help (indicated by CD4+ AIM and induction of Th1 cytokines), whereas T cell production of IL-4 and IL-10 were negatively associated with antiviral antibody functions (**Figure 5E**). The univariate correlation analysis between these T cell parameters, neutralization, and antibody effector functions in SARS-CoV-2 infected subjects further highlights that specific CD4+ T cell responses track with development of qualitatively different antibody responses (**Figure S6F**). Prior exposure to seasonal coronaviruses may also play a role in the development of qualitatively different antibodies as IgM responses against the seasonal coronaviruses, particularly 229E, were negatively associated with IL-4 and IL-10 (**Figure 5E**), reflective of our t-SNE analyses (**Figure 5C**).

Together, these data suggest that the balance of Th1/Th2 CD4+ T-cell responses played a key role in developing qualitative SARS-CoV-2 antibody responses and points to a potential role for IgM, but not IgG1, responses against seasonal coronaviruses in promoting Th1 immunity.

### Post-vaccination responses are predicted by Th1 immunity

With ongoing vaccinations, it is also unclear how prior infection will shape long-term and vaccine-mediated immunity. Vaccine responses and efficacy have been tied to the development of neutralizing antibody responses [11], although there is also strong evidence for T-cell or Th1 responses post-vaccination [8][35].

To understand how prior SARS-CoV-2 immunity resulting from infection impacted vaccine-mediated immunity, we obtained follow-up samples from a subset of infected subjects who received SARS-CoV-2 vaccine (n=32; age 16–79 years old (mean 43) and 63% female). These subjects received either the two-dose mRNA vaccines from Pfizer (n=21) or Moderna (n=11) or the single-dose J&J adenovirus vector vaccine (n=1). Blood collections spanned 2–315 days post-primary immunization, with 12 subjects having two or more samples collected (**Figure 6A**). As neutralizing antibodies have been linked to vaccine efficacy [20, 21], we determined post-vaccination neutralizing antibody titers. Comparison of neutralizing antibody levels pre- and post-vaccination demonstrated that vaccination significantly increased levels of neutralizing antibodies (**Figure 6B**). Waning immunity was evident over time post-vaccination, particularly 6 months following the second mRNA immunization (**Figure 6B-C**), as others have reported [21, 36, 37].

**Figure 6.**
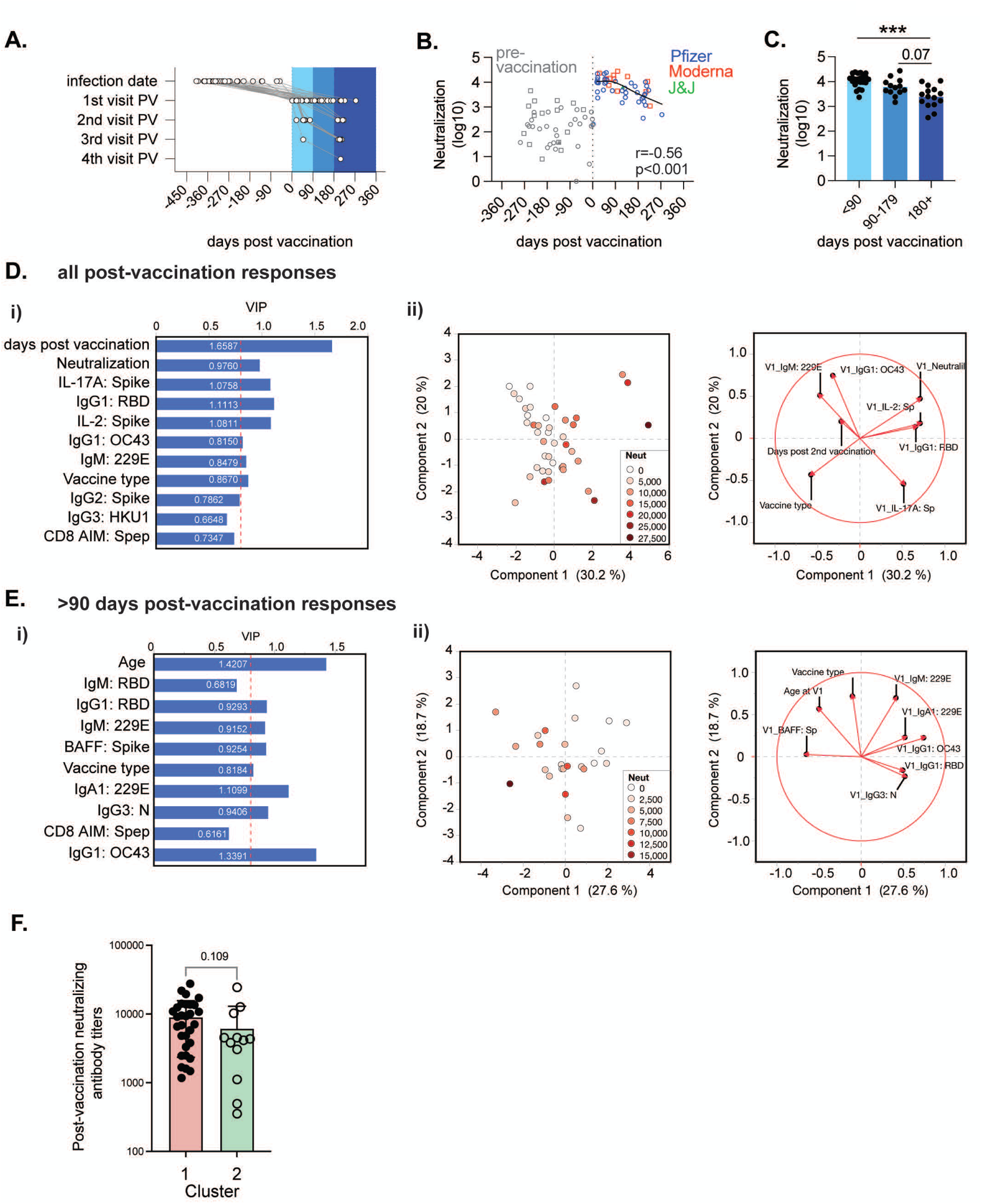
Post-vaccination responses are predicted by SARS-CoV-2 immunity post-infection. (A) Sample timeline for select subjects with SARS-CoV-2 infection who were followed for post-vaccination (PV) sample collections (n=32). (B) Neutralization antibodies in these subjects pre- or post-initiation of vaccination colored by vaccine type. Spline trend line (black) shown for post-vaccination samples with Spearman’s correlation indicated (for samples collected post-17 days). (C) Titers of neutralizing antibodies are plotted according to days from the completion of the primary vaccine series. (D-E) Predictive modeling with feature reduction analysis was used to determine the post-infection immune features that predicted neutralizing antibody titers across all post-vaccination time points (D) or >3 months (E). The selected features that best predicted post-vaccine neutralizing antibody titers in a partial least squares regression analysis are shown in (i), and features with a VIP score >0.8 were used in a principal component analysis (ii). (F) Post-vaccination neutralizing antibody titers in samples from subjects from cluster 1 or cluster 2. Mann-Whitney analysis was used to determine statistical significance as indicated or P <0.001 (***).

To determine if post-infection immune responses could predict neutralizing antibody responses to vaccination and help validate the importance of the t-SNE and network analyses in **Figure 5**, we performed a LASSO-selected regression analysis on neutralizing antibody responses observed across all subjects and time post-vaccination, or <90 days or > 90 days post-vaccination to identify the minimal features needed to predict neutralizing antibody responses within a given time frame. LASSO-selected features were then used in a partial least squares analysis to identify the variable importance in projection (VIP) features that predicted neutralization titers. Features with VIP scores >0.8 were then used to generate a PCA to determine those features that influenced vaccine-induced neutralizing antibody titer levels (**Figure 6D-E**).

Across all samples where collection time points spanned 2 to 315 days post-vaccination, one of the main driving factors that predicted higher levels of neutralizing antibodies was the number of days post-vaccination the samples were collected (**Figure 6D**), with samples collected before 90 days of vaccination having higher neutralizing antibodies than those collected afterwards. Post-infection immune parameters that predicted higher levels of neutralizing antibodies were CD4+ T cell production of IL-2, IL-17A, as well as the levels of IgG1 specific for RBD and neutralizing antibody titers post-infection (**Figure 6D**), suggesting that robust immune activation following infection translated to elevated immunity following vaccination. Curiously, CD8+ activation responses were negative predictors of post-vaccination neutralization responses (**Figure 6D-E, Figure S7**). The importance of pre-existing immunity to seasonal coronaviruses was also verified in this cohort as IgG1 OC43, IgM 229E, and IgG3 HKU1 were negative predictors of post-vaccination responses. Similar responses were observed within the samples collected within 3 months of vaccination and by correlation analyses (**Figure S7A-B**).

For samples collected after 3 months, older age, type of vaccine received, B-cell activating factor (BAFF), and lower levels of antibodies against the seasonal coronaviruses, including IgG1 OC43, IgM 229E, and IgA1 229E tracked with higher neutralizing antibodies (**Figure 6E, Figure S7B)**, suggesting that durable vaccine-mediated immunity is also shaped by prior SARS-CoV-2 and seasonal coronavirus immunity and age.

Finally, a comparison of the post-vaccination neutralizing antibody titers between subjects in population 1 (Th1 biased immunity) and population 2 (Th2 biased immunity) showed a trend towards higher neutralizing antibodies in population one post-vaccination (**Figure 6F**). This finding indicates that the two observed clusters partially explain diversity in SARS-CoV-2 antigen responses post-infection or vaccination based on CD4+ T-cell responses, age, and non-IgG1 antibody isotypes to SARS-CoV-2 or seasonal coronaviruses.

## DISCUSSION

This is among the first studies to comprehensively measure serological and cellular measures, combining a systems serology approach with T-cell and cytokine analysis. Our key findings confirmed that CD4+ Th1 responses and age track with qualitative features of humoral immunity, and we demonstrated that post-infection responses shape levels of vaccine-induced neutralizing antibody titers. Importantly, this study was performed in a non-hospitalized community cohort, encompassing individuals across a range of ages who experienced typical asymptomatic or mild-moderate disease, reflective of the vast majority of SARS-CoV-2 infections. As previously reported[10], infected subjects developed anti-spike, RBD, and nucleoprotein antibodies that exhibit polyfunctionality with multiple antiviral effector functions (**Figure 2, Figure S3**). Accordingly, CD4+ and CD8+ T-cell responses to spike and peptide pools were also significantly altered between infected and non-infected subjects, including high levels of Th1 cytokine expression characterized by high levels of IL-2 and also INFγ (**Figure 3**) as expected from previous studies [4, 24-28, 38].

We observed diversity in immune responses following infection in this cohort, including infected subjects that did not develop neutralizing antibody responses (**Figure 2**) and T-cell responses in the absence of seroconversion (**Figure S3**). Age but not sex correlated to many of our immune measures, BMI and to the symptom severity experienced by infected subjects (**Figure 4**). This would confirm that age is a primary driver of immunity, with reduced IgG and neutralizing antibodies observed in younger ages as expected from previous reports [39, 40]. However, other antibody functions, including complement and NK activation were unaffected. Curiously IgM and IgA levels did not correlate with post-infection duration and were not reduced over a second study visit (**Figure 2, Figure S2**), though older ages were associated with higher levels of IgM (**Figure 4**). IgM is traditionally an early response to infection, but sustained expression has been reported in asymptomatic or symptomatic convalescent patients [17, 41]; though others have found persistent IgM levels associated with reinfection or prolonged shedding of virus [42] or observed decreases immediately following infection [38]. We were unable to assess early IgM responses following infection, as the average study visit for this cohort was 93 days post-infection (**Table1**). Taken together the results presented here suggest that age-related responses to SARS-CoV-infection are likely related to isotypes generated during infection, altering antibody effector functions. Since IgM-expressing B-cells play important roles in humoral immunity for early and late antibody responses, and antigen presentation are associated with complement deposition [43, 44], it is possible that IgM levels in our cohort could be more than a temporal antibody maturation response and should be studied further.

The combined induction of functional cellular and humoral immunity is likely a key factor in long-term immunity. We observed a much higher correlation of antibody measures to CD4+ T-cells and cytokines than to CD8+ T-cells (**Figure 3, 5**), consistent with other reports on humoral memory [45, 46]. However, CD8+ T cell activation was also significantly detected and altered by age (**Figure 3, Figure 4**), and CD8 activation to spike peptide pools seemed to limit post-vaccination neutralization responses (**Figure 6**). This finding may reflect that specific CD8+ T-cell populations limit longevity of neutralization titers after infection, as others have suggested [47] or reflect a robust CD8+ T-cell response that is also protective [48]. Interestingly, combined analyses showed that infected subjects could be delineated into two groups based on a Th1-versus Th2-biased immunity that further tracked with differential development of neutralizing antibodies following vaccination. Importantly, individuals who developed a more Th1-biased immune response following infection, including production of Th1 cytokines from activated T-cells and development of antiviral antibodies capable of inducing both neutralization and innate immune effector function, had elevated levels of neutralizing antibodies post-vaccination (**Figures 5-6**). Antibody-dependent, complement-mediated delivery of antigen to dendritic cells has been shown to enhance production of neutralizing antibodies [43], thus in addition to having higher numbers of memory B cells, one potential hypothesis is that enhanced delivery of vaccine antigens to antigen presenting cells by existing SARS-CoV-2 specific antibodies helps drive higher neutralizing antibody levels.

The development of Th1-biased responses in subjects within cluster 1 tracked with overall older age, but not with symptom severity or SARS-CoV-2 infection. Intriguingly, subjects in cluster 2 reported experiencing more minor symptoms (e.g., nausea, runny nose (coryza), nasal congestion, cough, and headache) or specific symptoms (e.g., loss of taste) and had elevated levels of virus-specific IgG2 and IgG4 compared with subjects in cluster 1. While virus-specific IgG1 and IgG3 were not different between the two clusters, antibody functionality and neutralization were significantly different, highlighting that antibody features beyond IgG subclasses may further drive differences in antiviral functionality between the two groups. Antibody glycosylation can further modify the induction of innate immune effector functions. While we did not evaluate antibody glycosylation in this study, several studies have found that severe SARS-CoV-2 infection in hospitalized or critically ill subjects is associated with increased antibody afucosylation, leading to increased inflammation and innate immune cell-mediated immunopathology [49-52].

This study also revealed an association with Th2 cytokines, including IL-4, IL-13, and IL-10, which inversely correlated to neutralization after infection but only weakly to post-vaccination neutralization (**Figure 2, Figure 6, Figure S7B**). A Th2 profile has also been implicated in severe COVID disease [31], though these patients were not in our study population. IL-10 was strongly correlated with IgA response to spike in our cohort, which was a key factor in lower post-vaccination neutralization titers and longevity >3 months (**Figure 6, Figure S7**). A similar IL-4/IL-13 cytokine profile by T-cells was associated with the development of memory IgA+ B cells and not neutralization in cases of mild or severe COVID-19 [45]. However, others have shown that IgA is dominant as an early neutralizing antibody response to SARS-CoV-2 infection in mucosal tissue and blood and correlates to IgG [53, 54]. mRNA vaccination is known for boosting high levels of neutralizing serum antibodies but not mucosal IgA responses even in previously infected individuals [55]. However, these findings suggest that IgA responses play a negative role in post-vaccination neutralizing antibody magnitude and duration and may be the result of IL-10 secretion that that should be explored further. Taken together, this data indicate a key role for cellular immunity and cytokine biases in infection outcomes and post-vaccination responses.

Whether prior exposure to seasonal coronaviruses shapes SARS-CoV-2 specific immunity has been a key question since the beginning of the pandemic. Pre-pandemic pediatric samples have been a higher level of cross-reactive IgM to SARS-CoV-2, compared with adults who have higher levels of IgG and IgA, presumably from these seasonal coronaviruses [56]. These pre-existing antibodies, particularly to OC43, are increased following infection but not vaccination [57-59], OC43 antibodies have been linked to antibody development post-infection and COVID-19 survival in hospitalized patients [58] and are boosted following infection but not vaccination [57]. While no differences were observed in this cohort with respect to levels of antibody isotypes to seasonal coronaviruses by infection status (**Figure 2, Figure S1**), there were significant correlations between antibodies to SARS-CoV-2 spike, and nucleoprotein and those to spike proteins of seasonal coronaviruses (**Figure 2F**). Age did not correlate with the magnitude of these seasonal coronavirus IgG antibodies (**Figure 4**). We also observed that OC43 and NL63 IgG levels correlated with activation of T cells with spike peptide pools, but not protein (**Figure 3**), indicating this peptide pool contained epitopes known to overlap with seasonal coronaviruses as previously reported [60] that was not observed with full protein.

Variable reduction analyses identified population clusters that differed significantly in their antibody isotypes to seasonal coronaviruses (IgG1, IgM, IgA in population 1 vs IgG2, IgG4 in population 2, **Figure 5, Figure S6, Table S2**), suggesting that these antibody responses, particularly IgM 229E and IgG1 OC43, may impact the development of SARS-CoV-2 immunity. IgM responses to 229E spike protein were negatively correlated with Th2 cytokines IL-4 and IL-10 post-infection (**Figure 5E**) and post-vaccination neutralization responses (**Figure 6D-E, Figure S7A-B**). IgG1 levels to OC43 spike protein negatively impacted neutralization responses within <3 months post-vaccination (**Figure 6D, Figure S7A-B**), but positively affected longevity of post-vaccination responses (**Figure 6E, Figure 7B**). Thus, we confirmed evidence that pre-existing to seasonal coronaviruses immunity shapes infection and vaccination responses to SARS-CoV-infection [57-59]. However, this effect differed by time post-vaccination and supported an important role for non-IgG1 isotypes (IgM, IgG3) to seasonal coronaviruses that warrants further study.

Study limitations primarily involved using SARS-CoV-2 infection to differentiate subjects rather than pre-pandemic samples. In addition, the assays were limited to peripheral blood samples and not tissue-specific responses, which included only effector functions to spike protein and cytokine secretion instead of T-cell subset analyses. Detection of secreted cytokines allowed a greater number of cytokines to be evaluated but prevented confirmation of cells producing cytokines as would be observed intracellular stained cytokines for specific T-cell populations. However, cytokines between spike or peptide pools were highly correlated (**Figure S5**), indicating T-cell production. Also, high expression of IL-2 has been routinely observed from CD4+ T-cell and not CD8+ T-cells after SARS-CoV-2 infection [28, 38]. In this study, IL-17A secretion was closely correlated to IL-2 and Th1 cytokine release after stimulation with protein or peptide pools, suggesting that IL-17A may be serving as a proxy for a Th1/Th17 subset, as identified in other post-vaccination studies [61] which should be more closely examined. Finally, while the critical role for age in SARS-CoV-2 immunity was validated, it remains an ongoing question of why children exhibit less severity with infection and how differences in qualitative features of immunity depend on patient age.

Our study used samples collected from subjects only shortly after the pandemic which will be difficult to perform as COVID subsides and vaccination rates increase in the general population. Our findings indicate that SARS-CoV-2 specific humoral responses and functions are likely shaped by age, disease severity, quality of CD4+ T cell help, IgM or other non-IgG1 antibody isotypes to seasonal coronaviruses. SARS-CoV-2 specific T-cell responses, particularly IL-2 secretion, can be their own correlate of protection. Further studies combining powerful systems serology tools with T-cell function measures with other cohorts, tissue types, and timeframes will continue to tease out how these factors contribute to the diversity in SARS-CoV-2 infection outcomes and duration of immunity, including risks of breakthrough infections.

## Data Availability

All data produced in the present study are available upon reasonable request to the authors.

## Acknowledgements

We sincerely thank the CSI study participants. Funding for this study was provided under NIH Project U54CA260581-01. Funding for CVM and SSD was supported by the Telomere Research Network, NIH Project 5U24AG066528 (SSD). The funders had no role in study design, data collection and analysis, decision to publish or preparation of the manuscript.

## Author Contributions

Conceptualization, E.B.N., B.M.G., J.E.R, J.S.S., M.L.D., S.S.D., K.J.Z; Methodology, E.B.N., B.M.G., J.E.R; Investigation, A.E.M., E.E., I.V.T., T.J.Y., S.B., S.C., D.H.E., A.R.S., S.N.H.,K.E.O., S.J.B., B.N. T.H.; Resources, M.L.D., C.Y.Z., I.V.T., C.V.M., S.E.C., A.A., A.E.S., M.J.G., N.D.F., E.J.E., A.D.P., S.J.L.,J.K.K.; Data Curation, E.B.N., A.E.M., E.E., J.G.S.; Visualization, E.B.N., B.M.G., J.G.S.; Writing – Original Draft, E.B.N., B.M.G., A.E.M, EE, J.G.S.; Writing – Review & Editing, J.E.R., J.S.S., K.J.Z., J.G.S., J.K.K.; Funding Acquisition, E.B.N., B.M.G., J.E.R, J.S.S.

## TABLE AND FIGURE LEGENDS

**Table S1. Study questionnaire on demographic and COVID-19 factors**.

**Figure S1.**
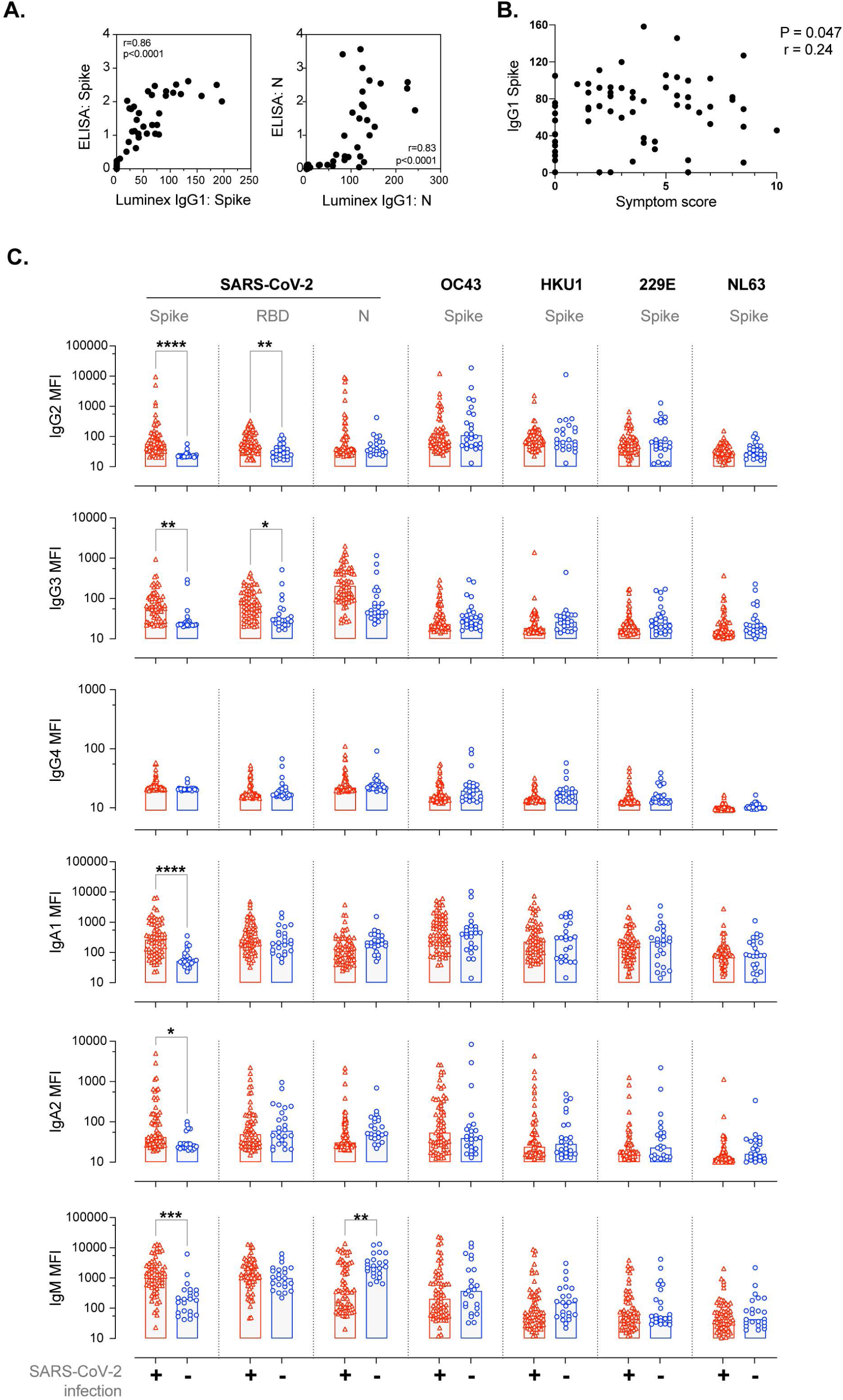
Expanded humoral analysis of the study population. (A) Strong concordance of results between spike and nucleoprotein IgG by Luminex-based analysis and ELISA optical density results. (B) Symptom score versus IgG1 responses to spike with Spearman’s correlation indicated. (C) IgG2, IgG3, IgG4, IgA1, IgA2, IgM responses to spike, RBD or N SARS-CoV-2 specific antigens, and spike-specific responses to seasonal coronaviruses OC43, HKU1, 229E, NL63 for each subject by SARS-CoV-2 infection status.

**Figure S2.**
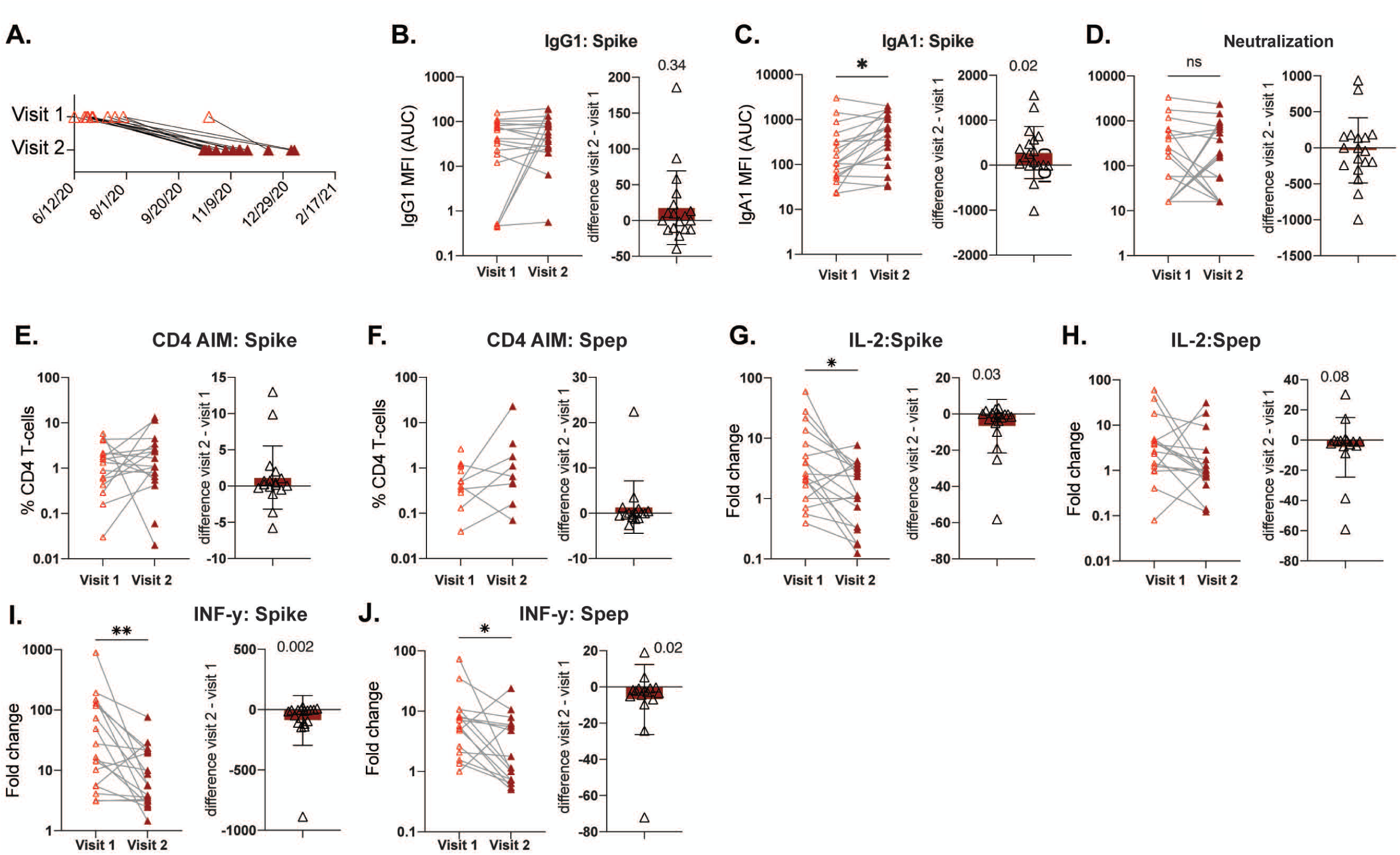
Stability of cellular and humoral measures between multiple study visits. (A) Schematic of original and return study visit by date for selected SARS-CoV-2 infected subjects (n=18). Immune measures were compared between first and second subject visits (indicated by line) and the difference calculated for each subject with bar at the mean +/- SD; including: (B) spike IgG1, (C) spike IgA1, (D) neutralization responses, (E) CD4 T cell AIM to spike, (F) CD4 T cell AIM to spike peptide, (G) IL-2 secretion to spike, (H) IL-2 secretion to spike peptide (fold change from unstimulated), (I) IFNγ secretion to spike, (J) IFNγ secretion to spike peptide. Significance by Wilcoxon signed rank test indicated as p<0.05(*), <0.01 (**), or <0.001 (***).

**Figure S3.**
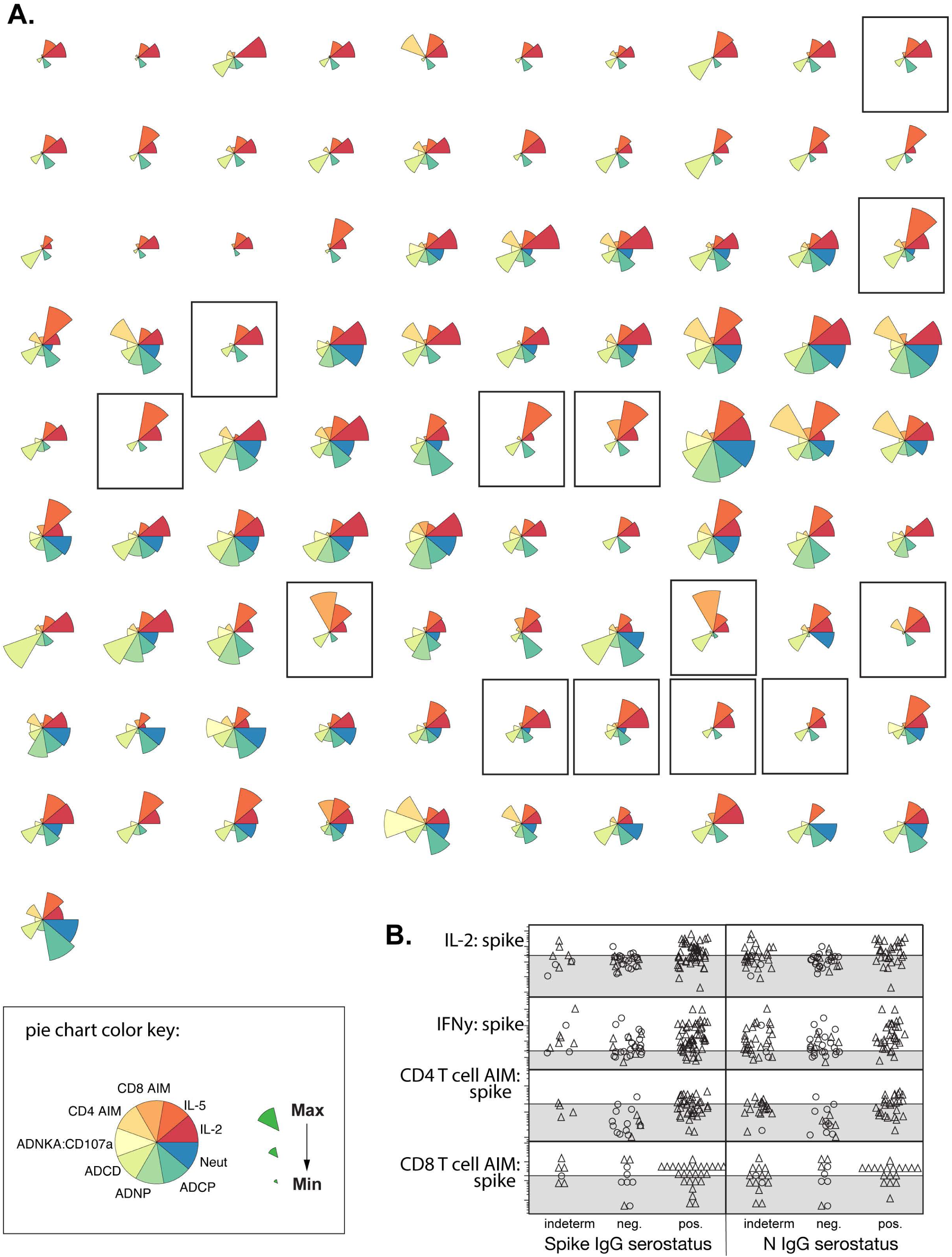
Individual subject responses. (A) Pie charts of each subject by antibody functions and select cellular measures. Black boxes indicate subjects with evidence for cellular immunity without seroconversion. (B) CD4+ T cell AIM, CD8+ T cell AIM, IL-2 secretion, or IFNγ secretion to spike protein by spike or nucleoprotein IgG serostatus.

**Figure S4.**
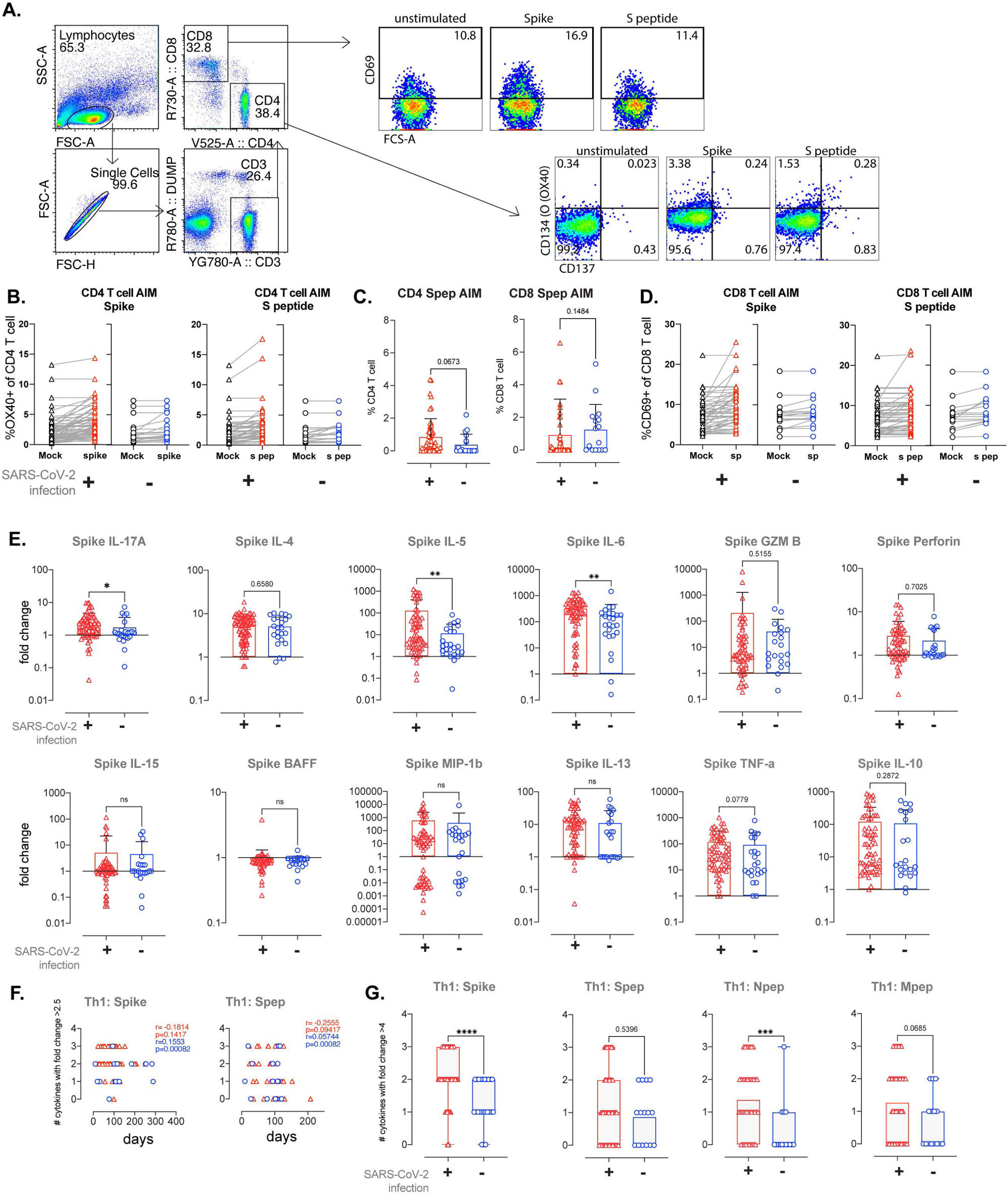
Measures of cellular immunity in study population. (A) Example AIM analysis by gating CD134-high expression on CD4 T-cells (CD4+, CD8-, CD3+ DUMP- (dead/CD33) or CD69 expression on CD8 T-cells (CD4-,CD8+,CD3+,DUMP-). (B) CD4 T cell AIM responses for mock and stimulated conditions for each subject (line) by infection status. (C) CD4 or CD8 T cells AIM to spike peptide pools by infection status. (D) CD8 T cell AIM responses for mock and stimulated conditions for each subject (line) by infection status. (E) Spike specific cytokine secretion expressed as a fold change from unstimulated cells by infection status. (F) Number of IL-2, IFNγ, or TNFα cytokines expressed at fold change >2.5 from unstimulated cells by day post infection or possible exposure. (G) Number of IL-2, IFNγ, or TNFα cytokines expressed with antigen stimulation at fold change >4 from unstimulated cells by infection status. Bars at mean +/- SD. Significance by Mann Whitney test indicated as P<0.05 (*), <0.01 (**), or <0.001 (***).

**Figure S5.**
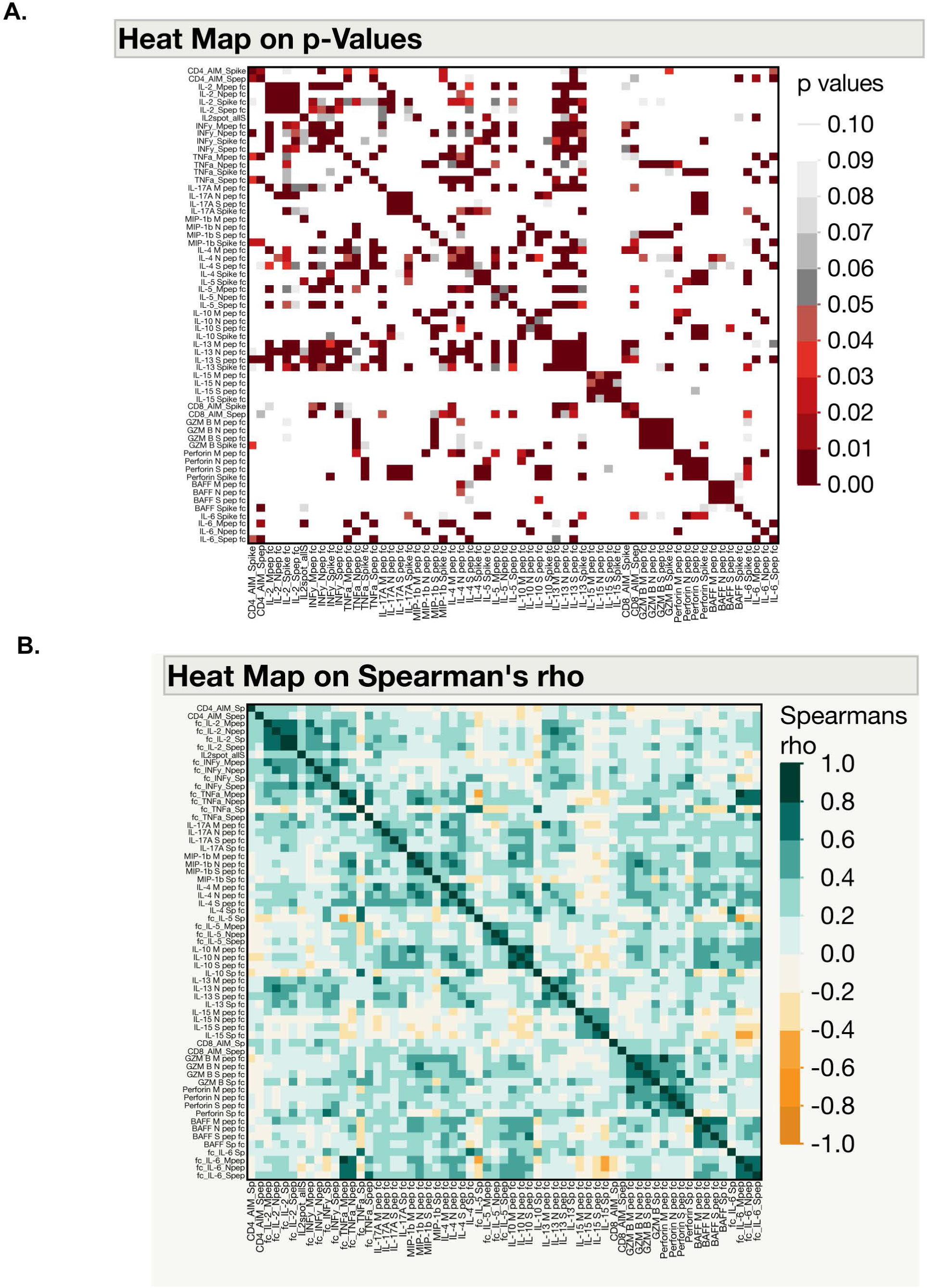
Strong correlation between stimulation antigens used for T-cell AIM and cytokine secretion assays. Correlation matrix for indicated measures after stimulation with spike protein or spike, nucleoprotein, or matrix peptide pools stimulation displayed as heatmaps of indicated Spearman’s p values (A) and rho correlation coefficient (B).

**Table S2. Kruskal-Wallis analyses comparing t-SNE Cluster 1 and 2 populations and demographic, infection, or immunologic variables**.

**Figure S6.**
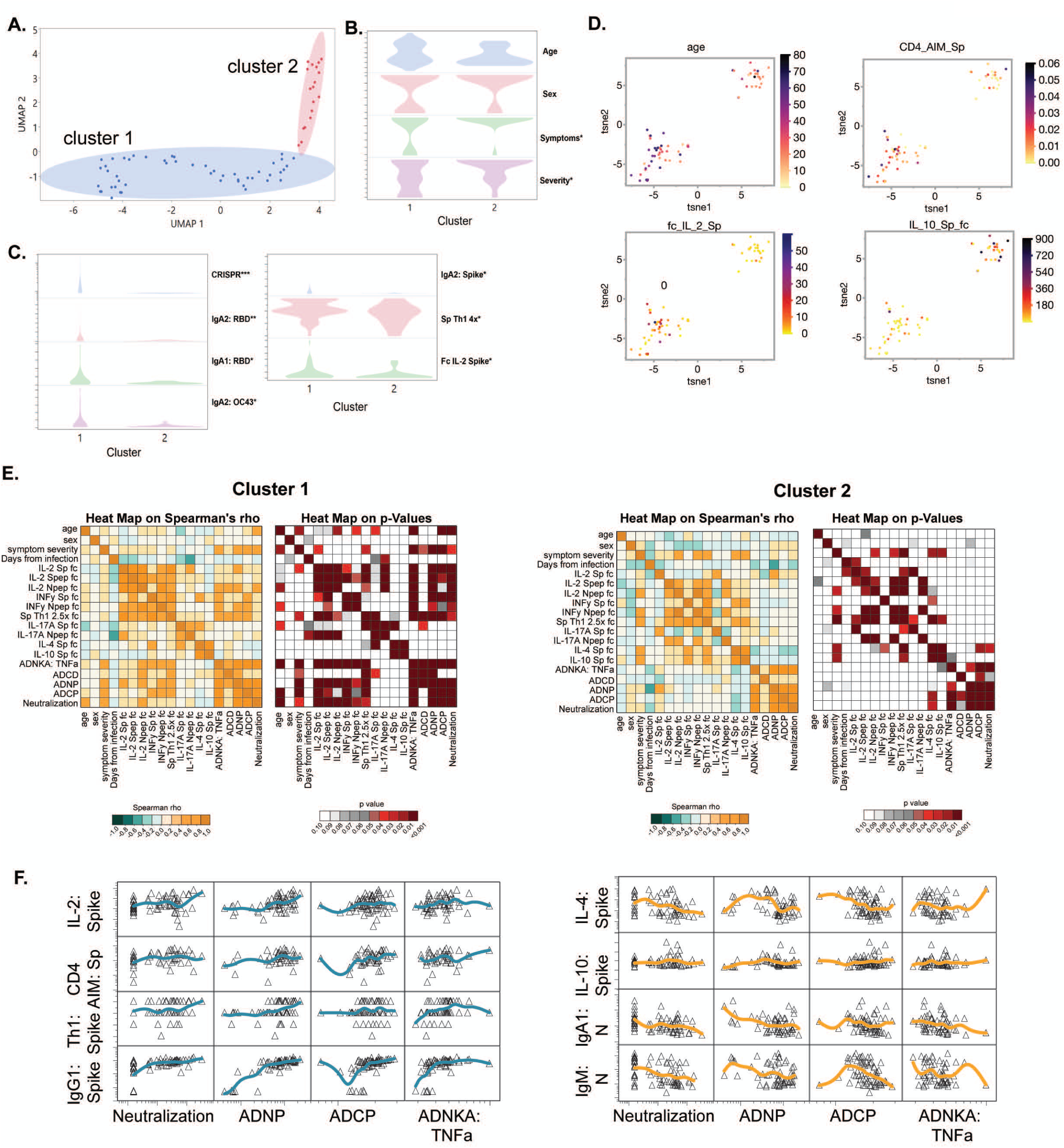
Supporting data for population clusters by SARS-CoV-2 immunity. (A) Imputed dataset for all SARS-Co-V immunologic variables was used for UMAP analysis. The blue and red shaded regions represent clusters of UMAP values determined by the K-means clustering approach. All imputed data were required to have at least 60% of non-missing cell values. (B) Graphs of demographic and SARS-CoV-2 infection variables with correlation significance to UMAP 1 or 2 indicated P<0.05 (*). (C) Graphs of variables (P<0.05) significantly correlated with UMAP 1 or 2 at P<0.05. (D) Additional tSNE analysis graphs color-coded with indicated variables for SARS-CoV-2 infected subjects only. (E) Correlation matrix for indicated measures with heatmaps of Spearman’s rho values and p values by cluster 1 (right) or cluster 2 (left). (F) Validation of key tSNE findings using unimputed dataset from SARS-CoV-2 infected subjects with spline trend line (blue or orange) shown all graphs in log scale except Th1 cytokines to Spike (number of >2.5 fold change for IL-2, IFN, TNF).

**Figure S7.**
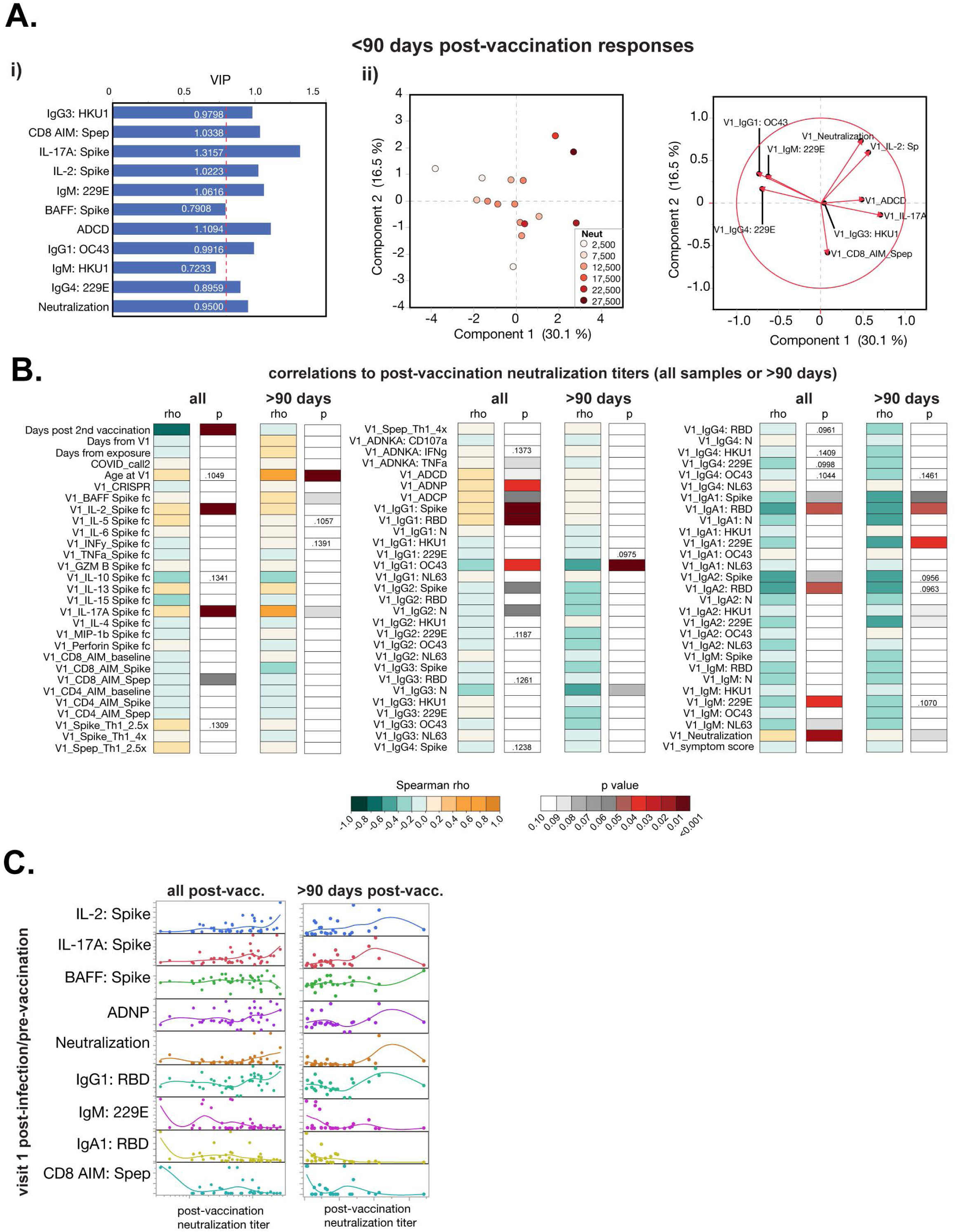
Expanded analysis of post-infection immune features that predict or correlate with neutralizing antibody titers after vaccination. (A) Predictive modeling with feature reduction analysis was used to determine if specific post-infection immune features predicted post-vaccine neutralizing antibody titers <3 months. The selected features that best predicted post-vaccine neutralizing antibody titers in a partial least squares regression analysis are shown in (i), and features with a VIP score >0.8 were used in a principal component analysis (ii). (B) Correlation matrix for post-infection/pre-infection immune measures to post-vaccination neutralization responses with heatmaps of indicated Spearman’s rho and p values for all samples or >90 days samples from completion of vaccination series. (C) Graphs of select measures from post-infection/pre-infection immune measures to post-vaccination neutralization responses for all samples or >90 days samples from completion of vaccination series.

## Methods

### Study design and subject recruitment

The study was a rolling prospective cohort of subjects recruited from communities in New Orleans between June 2020 to February 2021. Subjects or households with suspected or confirmed SARS-CoV-2 infection were recruited from the Greater New Orleans community under Tulane Biomedical Institutional Review Board (federalwide assurance number FWA00002055, under study number 2020-585). Enrolled subjects completed a study questionnaire regarding infection and demographic information and provided a blood sample. A subsample of subjects returned for a follow-up visit.

### SARS-CoV-2 infection and COVID-19 case classification

History of SARS-CoV-2 infection was defined as 1) clear evidence of immunity (SARS-CoV-2 S or N-specific IgG), or 2) detection of plasma viral RNA as described below, or 3) fulfillment of the Centers for Disease Control and Prevention (CDC) case definition of confirmed or probable COVID-19 infection (an individual with a] confirmatory or presumptive laboratory criteria including history of positive SARS-CoV-2 PCR or antigen test or b] absence of negative PCR/antigen test that was performed 2 days prior to 5 days after onset of symptoms and clinical criteria with certain symptoms and fulfill the epidemiological criteria with exposure to a family or household contact with known SARS-CoV-2 [62]).

### Symptoms and illness severity scores

A general symptom score was calculated according to the September 2021 CDC classification (prior to circulation of the Omicron variant). Two points were given according to each major symptom: cough, shortness of breath, difficulty breathing, loss of smell, and loss of taste. One-half of a point was given for each minor symptom: fever, runny nose, nasal congestion, sore throat, diarrhea, vomiting, fatigue, muscle aches, chills, nausea, and headache. Total symptom score was calculated as the sum of the major and minor scores.

Because illness severity is not necessarily measurable by the point-based approach used for the general symptom score, it was calculated according to clinical manifestations thresholds based on lung involvement and impairments, as detailed in Baj et al. (2020, [63]). Specifically, the severity score was defined as severe (abdominal pain, shortness of breath, difficulty breathing, chest pain, vomiting, nausea, diarrhea); moderate (fever, cough, runny nose, fatigue, muscle aches, loss of smell, headache, sore throat, loss of taste, nasal congestion, chills, repeated chills, hair loss, toes); and asymptomatic (no reported symptoms). Hair loss was considered to be typically associated with long-term COVID-19 symptoms and thus was not considered as an acute, severe symptom.

### Continuous predictors

Age was based on self-reported birth date at study enrollment. Body mass index (BMI) was defined as weight in kilograms divided by height in meters squared. For adults, BMI was calculated as: as <18.5 (underweight), 18.5-30 (normal), and >30 (overweight/obese). For children (aged under 18 years), BMI was calculated as <5^th^ percentile (underweight), between the 5^th^ and 95^th^ percentile (normal), and greater than the 95^th^ percentile (overweight/obese).

### Sample collections

Blood was collected from subjects and plasma and peripheral blood mononuclear cells (PBMCs) and were isolated by density gradient centrifugation in Leukosep tubes (Greiner Bio One) and Ficol-Paque PREMIUM 1.078g/ml (Cytiva) [64, 65]. Plasma was removed and stored at -80°C or heat-inactivated at 56°C for 30mn before testing. PBMC were washed, counted, and suspended in FBS-10% DMSO at 1×10^7^ cells/ml. Aliquots of cells were frozen at -80C in a Nalgene Mr. Frosty container (Nalgene Labware, Rochester, NY) before final storage in liquid nitrogen.

### Plasma viral RNA detection

RNA viral load was measured with a sensitive SARS-CoV-2 specific CRISPR assay as previously reported [33, 34]. Twice the limit of viral detection, or 2×10^6^ as RNA abundance expressed as the relative photoluminescence intensity of the sample was used as definitive evidence of SARS-CoV-2 infection.

### Determination of antigen-specific antibody reactivity by multiplexed Luminex analysis

Recombinant SARS-CoV-2 antigens (full-length spike, RBD, and N) and the recombinant spike protein from OC43, HKU1, 229E, and NL63 (Frederick National Laboratories) were coupled with MagPlex beads (Luminex) via sulfo-NHS coupling chemistry. Heat-inactivated samples were diluted at 1:50 in 1X PBS + 0.1% bovine serum albumin (BSA) + 0.05% Tween20 and incubated with antigen-coupled beads for 2 hours. Beads were washed and incubated with 0.65µg/ml of PE-labeled secondary antibodies specific for the human antibody subclasses IgG1 (Clone HP6001; Southern Biotech), IgG2 (Clone HP6002; Southern Biotech), IgG3 (Clone HP6050; Southern Biotech), IgG4 (Clone HP6025; Southern Biotech), IgA1 (Clone B3506B4; Southern Biotech) IgA2 (Clone A9604D2; Southern Biotech), IgM (Clone UHB, Southern Biotech) for 1 hour at room temperature. Beads were washed three times with assay buffer and analyzed on a MagPix instrument (Luminex, Austin, TX). The median fluorescent intensity for 50 beads/region was recorded. SARS-CoV-2 seronegative plasma samples were used to establish baseline reactivity and thresholds for positivity.

### ELISA and Neutralization assays

Recombinant SARS-CoV-2 spike for use in ELISAs was produced in-house by stable expression of a modified phCMV based plasmid that encodes the pre-fusion trimeric SARS CoV-2 spike protein with the D614G mutation (kindly provided by Dr. Kate Hastie (La Jolla Institute for Immunology)[66, 67]). Stable expression was maintained via blasticidin selection in stably transfected 293F cells. Recombinant protein was purified from clarified culture medium by affinity chromatography on Streptactin X affinity columns. RBD (aa321-535) was similarly expressed in the phCMV plasmid and purified on Streptactin X affinity columns.

A DNA fragment encoding SARS CoV-2 N protein, including its natural leader sequence was generated by PCR of full-length N protein gene from a lentiviral N Protein expression vector (pLVX-EF1alpha-SARS-CoV-2-N-2xStrep-IRES-Puro, which was a gift from Nevan Krogan (Addgene plasmid # 141391 ; http://n2t.net/addgene:141391; RRID:Addgene_141391, [68]). This fragment was inserted in place of the RBD sequence in the above expression plasmid upstream of the double strep-tag sequence using NEB gene builder.

Wells of 96-well ELISA plates (Costar, Easy Wash) were coated for 1 hour at room temperature with purified SARS CoV-2 spike protein (500 ng/ml in 100 mM sodium bicarbonate buffer). Wells were washed X 5 and blocked for 1 hour at +37°C. Blocking and dilution buffers consisted of 0.5%Tween +5% dry milk+ 4%whey protein (BiPro, Le Sueur, MN) +10%FBS in 1xPBS. After wells had been washed and blocked, 100 ul heat-inactivated sera or plasma samples diluted to 1:100 were incubated in antigen-coated and uncoated wells for 1 hour. Wash buffer consisted of PBS with 500 mM NaCl and 0.2% Triton X. The higher salt content of the wash buffer greatly reduced background noise. Bound IgG was detected with peroxidase-conjugated goat anti-IgG (Jackson ImmunoResearch) and diluted at 1:15,000. Color was developed with TMB-H202 and stopped with 1 M phosphoric acid. Absorbance (optical density [OD]) was read at 450 nm. In experiments where samples were tested in wells with and without spike protein, net ODs were calculated by subtracting background OD readings from OD readings with spike protein. Cut-off OD values were then calculated based on testing of >100 pre-COVID-19 samples. Net OD values >0.5 were considered positive. The Spike protein ELISA for IgG antibodies has been validated by testing a standard set of positive and negative samples provided by NCI SeroNet staff. These validations showed sensitivity and specificity for the imunoassay as 98% and 100%, respectively).

Similar ELISAs were run with purified RBD and N protein-coated in wells. For the RBD ELISA, protein was coated at 500 ng/ml in 100 ul of 100 mM sodium bicarbonate. For the N protein ELISA, protein was coated at 2 ug/ml. Otherwise, the ELISA procedure was the same as for the spike protein ELISA.

### Neutralization of SARS CoV-2 in Pseudovirus Assay

CHO cells were generated and stably expressed ACE2 by transfecting CHO cells with an ACE2 expression plasmid containing the blasticidin resistance gene. ACE2 expressing cells were selected in medium containing 10 ug/ml blasticidin ml. Blasticidin-resistant cells were expanded and selected for high level ACE2 expression by flow cytometry of cells binding a FITC-labeled murine Mab to ACE2 (Sino Biologicals Cat # 10108-MM37-F). ACE2 positive cells have been sorted twice and have stably expressed ACE2 through multiple passages over 4–5 months with gradual diminishment of luciferase signal induced by pseudovirus infection. Cell lines were cryopreserved, and, as needed, cells were periodically thawed and freshly grown for continued studies. CHO-ACE2 cells were similar in SARS CoV-2 susceptibility to the 293T/ACE2 cell line developed by Dr. Farzan [69] but have better adherence to tissue culture surfaces.

SARS-CoV-2 neutralizing antibodies were assessed in serum or plasma samples with sensitive, high-throughput pseudovirus assays. Virus neutralization was measured in CHO/ACE2 cells. The pseudovirus assay was originally developed by Drs. Barney Graham and others at Vaccine Research Center at the National Institutes of Health (NIH). The assay was formally optimized and validated in Dr. David Montefiori’s laboratory at Duke University. The materials and protocol were kindly provided to the Robinson lab by Drs. Graham and Montefiori.

For pseudovirus production, four expression plasmids were obtained from NIH Vaccine Research Center. These included an expression plasmid for full-length spike protein of the Wuhan-1 strain containing the D614G amino acid chain (VRC7480.G614) [70], a pCMV ΔR8.2 lentivirus backbone plasmid (VRC5602) [71], the VRC5601 plasmid pHR’ CMV Luc containing the firefly luciferase reporter gene [71], and VRC9260 for TMPRSS2 expression. Pseudoviruses were produced by co-transfection of the four plasmids into 293T cells grown in T75 flasks with Fugene 6 as transfection reagent. Virus stocks are collected 3 days after transfection, clarified, passed through a 0.45 μm filter, and stored in aliquots at -80°C.

For neutralization, a predetermined optimal dose of pseudovirus was incubated with 8 serial 3-fold dilutions of heat-inactivated serum or plasma samples in 150 μl medium for 1 h at 37°C in 96-well tissue culture plates. CHO/ACE2 cells, suspended by the action of TrypLE enzyme, were added to wells (10,000 cells in 100 μL medium per well). One set of eight control wells received cells + virus (virus control), and another set of eight wells received cells only (background control). After 66-72 hours of incubation, the medium was removed by gentle aspiration. Then, 100 ul of 1:6 dilution of Promega BriteGlo in Glo lysis buffer was added to the wells with mixing. Plates were incubated for 7 minutes at room temperature, after which luminescence was measured in a Biotek Synergy H1 Luminometer. Neutralization titers were defined as the serum dilution (ID50) at which relative luminescence units (RLU) were reduced by 50% compared to virus control wells after subtraction of background RLUs (determined by GraphPad Prism, version 9 for macOS, GraphPad Software, San Diego, California USA).

### Determination of Spike and Nucleoprotein IgG seropositivity

ELISA and Luminex results were compared for Spike or Nucleoprotein seropositivity for IgG or IgG1, respectively. Any consistent results, disagreements or borderline positive/negative results were coded as indeterminant.

### Antibody-dependent NK cell degranulation and activation

NK92 cells (ATCC CRL-2407) expressing CD16 were cultured in alphaMEM (Gibco; Cat# 12000-022) supplemented with 12.5% FBS (Hyclone SH30071.03), 12.5% horse serum (Hyclone; Cat# SH30074.03), 1.5g/L sodium bicarbonate (Fisher Sci; Cat# S-233-500), 0.02 mM Folic acid (Alfa Aesar; Cat# J62937), 0.2 mM Inositol (MP Biomedical; Cat# 194688), 0.1 mM Beta-mercaptoethanol (Gibco; Cat# 21985-023), 100 U/ml recombinant IL-2 (StemCell; Cat# 78036.3). Recombinant SARS-CoV-2 spike was coated onto MaxiSorp 96-well plates (Thermo Scientific Cat# 442404) at 300 ng/well at 4°C overnight. Wells were washed with PBS and blocked with 5% BSA. Serum samples diluted 1:50 in PBS were added to the wells and incubated for 2 h at 37 °C. Unbound antibodies were removed by washing wells with PBS. NK92 cells in complete alphaMEM culture medium were added at 5 × 10^4^ cells/well in the presence of 4 µg/ml brefeldin A (Biolegend Cat# 420601), 5 µg/ml GolgiStop (BD Biosciences Cat# 554724) and 0.15µg of anti-CD107a antibody (Clone H4A3 PE-Cy7, Biolegend Cat# 328618) for 5 hours at 37°C. Cells were stained for surface expression of CD16 (Clone 3G8 Pacific Blue, Biolegend Cat# 302032) and CD56 (clone 5.1H11 AlexaFluor488 Biolegend, Cat# 362518). Cells were fixed and permeabilized with Fix/Perm (Biolegend Cat# 421002) according to the manufacturer’s instructions and stained for intracellular IFNg (Clone B27 PE, Biolegend Cat# 506507), and TNFa (clone Mab11 APC, Biolegend Cat# 502912). Cells were analyzed on a Cytek Aurora spectral flow cytometer.

### Antibody-dependent neutrophil phagocytosis (ADNP)

Protocol was adapted from [72]. Recombinant biotinylated SARS-CoV-2 spike protein was coupled to Neutravidin fluorescent beads (LifeTechnologies). Serum samples diluted 1:100 in culture medium were incubated with spike-coated beads for 2h at 37°C. Freshly isolated white blood cells from human donor peripheral blood (5×104 cells/well) were added to wells and incubated for 1 hour at 37°C. Cells were stained for CD66b (Clone G10F5; Biolegend), CD3 (Clone UCHT1; BD Biosciences), and CD14 (Clone MjP9; BD Biosciences), fixed with 4% paraformaldehyde, and analyzed by flow cytometry on a Cytek Aurora spectral flow cytometer. Neutrophils were defined as SSC-Ahigh CD66b+, CD3neg, CD14neg. A phagocytic score was determined using the following formula: (percentage of bead+ cells)*(geometric mean fluorescent intensity (gMFI) of red bead+ cells)/10,000.

### Antibody-dependent cellular phagocytosis by human monocytes (ADCP)

Protocol was adapted from [73]. THP-1 monocytes were maintained in RPMI-1640 supplemented with 10% FBS, penicillin/streptomycin (Gibco Cat#15070063), L-glutamine, and b-mercaptoethanol. Recombinant SARS-CoV-2 spike-coated beads were generated as described for ADNP. Serum samples were diluted in a five-fold dilution curve in THP-1 culture medium (1:2500, 1:12500, 1:62500) and incubated with spike-coated beads for 2 h at 37 °C. Unbound antibodies were removed by centrifugation before adding THP-1 cells at 2.5×10^4^ cells/well. Cells were fixed with 4% paraformaldehyde and analyzed by flow cytometry for uptake of fluorescent beads on a Cytek Aurora spectral flow cytometer. A phagocytic score was determined as described above, and the areas under the curve for the three dilutions were calculated.

### Antibody-mediated complement deposition (ADCD)

Protocol was adapted from [74]. Recombinant SARS-CoV-2 spike-coated beads were generated as described for ADNP. Serum samples were diluted in culture medium 1:10 and incubated with spike-coated beads for 2 hours at 37 °C. Unbound antibodies were removed by centrifugation prior to the addition of reconstituted guinea pig complement (Cedarlane Labs CL4051) diluted in veronal buffer supplemented with calcium and magnesium (Boston Bioproducts) for 20 min at 37°C. Beads were washed with PBS containing 15 mM EDTA and stained with an FITC-conjugated anti-guinea pig C3 antibody (MP Biomedicals). C3 deposition onto beads was analyzed by flow cytometry on a Cytek Aurora spectral flow cytometer. The gMFI of FITC of all beads was measured.

### PBMC restimulation and culture

6-7.2×10^5^ PBMCs were cultured for 24 h in the presence of Spike Glycoprotein 0.2μg/well (BEI, NR-52308, produced under HHSN272201400008C and obtained through BEI Resources, NIAID, NIH: Spike Glycoprotein (stabilized) from SARS-Related Coronavirus 2, Wuhan-Hu-1 with C-Terminal Histidine Tag, Recombinant from Baculovirus), and SARS-CoV-2 specific mega pools at 0.2 μg/well including PepTivator SARS-CoV-2 Prot_S (Miltinyi - 130-126-700), SARS-CoV-2 Prot_M (130-126-702), SARS-CoV-2 Prot_N (130-126-699) in 96-well U bottom tissue culture plate (CytoOne CC7672-7596) in 200 μl RPMI-1640 with 10% FBS. Media was used as a negative control. Supernatants were harvested at 24 h post-stimulation for multiplex cytokines detection and stored at -80°C.

### Activation-induced marker (AIM) staining & detection

Restimulated cells from above were pelleted and stained with Fixable Viability Dye eFluor™ 780 in DPBS (eBioscience) for 30 minutes at 4C in the dark, then washed and resuspended in 50 μl with an antibody cocktail in Brilliant Stain Buffer (Biosciences) [75, 76] for 30 minutes at 4C. Cells were then washed with flow buffer (500 ml 1x PBS ThermoFisher 20012-027, 5 g BSA Roche 10735078001, 0.5 g Sodium Azide Sigma S2002) and fixed for 20 minutes at 4C with 10% formalin (polyScience 04018-1). Following fixation, cells were washed and resuspended in 200 μl flow buffers. All samples were acquired on a BD LSRFortessa. A list of antibodies used in this panel can be found table 1 below:

**Table 1.** Cellular antibodies used for AIM assay.

| <u>Stain</u> | <u>Source</u> | <u>Catalog number</u> | <u>Antibody</u> |
| --- | --- | --- | --- |
| FITC | BioLegend | 356914 | anti-CD185 |
| AF700 | BioLegend | 344724 | anti-CD8 |
| PE-Cy7 | BioLegend | 300914 | anti-CD8a |
| BV510 | BioLegend | 317444 | anti-CD4 |
| FITC | BioLegend | 310904 | anti-CD69 |
| APC-Cy7 | BioLegend | 366614 | anti-CD33 |
| PerCP-Cy5.5 | Invitrogen | 45-0338-42 | anti-CD33 |
| PE | BioLegend | 350004 | anti-CD134 |
| PE-Cy7 | BioLegend | 300420 | anti-CD3 |
| eFlour450 | Invitrogen | 48-0037-42 | anti-CD3 |
| APC | BioLegend | 309810 | anti-CD137 |
| APC-CY7 | ThermoFisher | 65-0865-14 | Live/Dead |

*Note.* All samples were acquired on a BD LSRFortessa.

### Cytokine testing

Cytokine test kits were used to measure cytokine levels in supernatant fluids from restimulated PBMC cultures according to manufacturers’ protocol. Kit types included Bio-plex Pro Human Cytokine 17-plex (Biorad) or custom human (Biorad Bio-plex IFN-y, IL-2, IL-5, IL-6, TNFα; Biolegend Legendplex Perforin, and Granzyme B, BAFF, MIP-1β, IL-4, IL-10, IL-13, IL-15, IL-17A). Samples tested with the Legend-plex kit were reanalyzed with the standards for each cytokine with the best range. Results from comparing cytokine levels in stimulated and mock stimulated cultures were expressed as fold change. This calculation reduced batch effects among experiments. Values of zero from bio-plex kits were replaced with 0.1 or 1 (TNF-α only) or for Legend-plex kits with 0.1 or 1 (MIP-1β and BAFF only).

### Statistical Analysis

Continuous data were expressed as means and standard deviations, medians and ranges, or medians and interquartile ranges. Categorical data were presented as frequencies and percentages. General comparisons among continuous predictors were performed using Krusal-Wallis test with Dunn’s post-tests for pairwise comparisons. General comparisons for categorical responses were performed using Pearson’s chi-square test (or Fisher’s Exact test where the assumption of a normal approximation to the binomial distribution was not justified). Pairwise comparisons for categorical responses were performed using logistic regression model contrasts. Dimensionality reduction was performed using t-Distributed Stochastic Neighbor Embedding [t-SNE] and Uniform Manifold Approximation and Projection [UMAP]) using the JMP Embedding Add On [77]. Feature selection using lasso regression was performed in JMP Pro 16. Lasso-selected features were used in a partial least square regression analysis with a leave-one-out validation method in JMP Pro 16. Features with a variable importance in projection score (VIP) >0.8 were used to generate a principal component analysis in JMP Pro 16. Data management and statistical analyses were carried out using the FloJo (version 10 Becton, Dickinson and Company, Ashland, OR), R (Version 4.1.2, R Foundation for Statistical Computing, Vienna, Austria) GraphPad Prism (version 9.0.0, GraphPad Software, San Diego, CA), JMP (version 16.2.0, SAS Institute, Inc., Cary, NC), and SAS (version 9.4, SAS Institute, Inc., Cary, NC). The type I error threshold was set at 5%.

### Data imputation for dimension reduction analyses

Data were singly imputed for tSNE and UMAP analyses using mean imputation for continuous variables and mode imputation for categorical variables. All imputed data were required to have at least 60% of non-missing cell values. Those variables not including at least 60% of observed data were excluded from the dimension reduction analyses.

